# A healthy donor kidney aging signature estimable from PAS histology and its association with progression of diabetic kidney disease

**DOI:** 10.64898/2026.11.05.26367347

**Authors:** Shaun G. Hong, Dongkwan Ryu, Hyunah Ku, Ara Ko, Jae-Ik Oh, Jinsun Lee, Min-Woo Kang, Myeongseop Kim, Youmin Shin, Hajeong Lee, Seung Seok Han, Yaerim Kim, Jinkyung Kwon, Jeonghwan Lee, Jungpyo Lee, Ji In Park, Jung Tak Park, Jung Nam An, Hyun Je Kim, Kyung-Chul Moon, Young-Gon Kim, Sehoon Park, Dong Ki Kim, KORNERSTONE investigators

## Abstract

**Background:** As kidneys age, their cells change in arrangement and gene expression. Whether these changes are also present in diabetic kidney disease (DKD), and whether they relate to how quickly kidney function is lost, is not known.

**Methods:** Zero-time biopsies from 24 healthy living kidney donors (aged 36–66 years) and biopsies from 28 patients with DKD were profiled by single-cell-level spatial transcriptomics with a 5,101-gene panel. In the donors, we identified how cell neighborhoods and cell-type gene expression change with age and summarized the expression changes as tissue-aging scores. The scores were applied unchanged to DKD and related to the rate of eGFR decline after biopsy in 24 patients, with adjustment for age, sex, baseline eGFR, and proteinuria. We also estimated the scores from routine periodic acid–Schiff (PAS) slides, using models that did not include the patient being scored.

**Results:** In older donors, immune cells were less often found next to tubular epithelium, and contacts between endothelial cells and podocytes and between stromal and epithelial cells were fewer, although cell proportions changed little. Expression of stress-response and cell-cycle regulators (*FOXO3*, *CDKN1A*, *BBC3*, *PINK1*, *IRAK1*, *SOCS1*) increased. Changes in cell contacts in DKD tissue largely followed the same direction as those in older donors. In DKD, higher tissue-aging scores were associated with faster eGFR loss independently of age, sex, baseline eGFR, and proteinuria (overall score, partial ρ = −0.61); this held for tubular, endothelial, immune, and parietal epithelial cell scores but not for the podocyte score. Scores estimated from PAS slides correlated with the transcriptomic scores (mean ρ = 0.62) and were also associated with faster eGFR loss (partial ρ, −0.44 to −0.56).

**Conclusions:** Healthy kidney aging involves a selective loss of cell contacts together with a coordinated stress-response program. DKD kidneys that carry this aging phenotype lose function faster, and the phenotype can be estimated in part from the routine diagnostic PAS slide.

## Introduction

The kidney ages in ways that can be seen in tissue [1]. Even in healthy adults, glomerulosclerosis, tubular atrophy, and interstitial fibrosis become more common with age [2, 3], and the age-related loss of kidney function largely reflects the loss of nephrons [4]. Aging kidneys also show a characteristic transcriptional profile [5], shaped in part by inflammatory transcription factors [6], with increased expression of cell-cycle and senescence-associated genes [7] and an altered extracellular matrix signature [8]. These changes vary between individuals, so kidneys of the same chronological age can differ in their biological state.

Whether this aging pattern is also present in diseased kidneys, and whether it affects their course, is less clear. The question is clinically relevant in diabetic kidney disease (DKD), in which patients of similar age and kidney function lose function at very different rates [9, 10]. Glomerular injury, tubular stress, inflammation, and interstitial remodeling drive DKD progression and overlap with changes seen in aging [11, 12]. A DKD kidney whose tissue resembles that of an older healthy kidney may therefore decline faster, and an aging signature defined in healthy kidneys could help explain why patients with similar clinical measures progress differently.

Answering this requires a method that keeps the arrangement of cells while measuring their gene expression. Single-cell-level spatial transcriptomics places transcripts from a large gene panel within individual cells, so that cell identity, expression, and neighboring cells can be read in the same section [13, 14]. Studies of human kidneys with single-cell and spatial methods have mapped injury states and disease-associated cell neighborhoods [15, 16], and spatial analysis can measure how often cell types lie next to one another [17]. These assays are not part of routine care, but every diagnostic kidney biopsy is examined with periodic acid–Schiff (PAS) staining, and pathology foundation models can infer tissue properties from such images [18]. The molecular state of a biopsy might therefore be estimated from its PAS slide.

Here, we used zero-time biopsies from healthy living donors to define how cellular neighborhoods and cell-type expression change with kidney age. We then asked whether this aging phenotype is present in DKD, whether DKD kidneys that carry it lose function faster, and whether it can be estimated from the PAS slide obtained in routine care.

## Methods

### Study cohorts

Participants were enrolled in KORNERSTONE, a prospective multicenter kidney biobank with linked clinical follow-up [19]. Spatial transcriptomic profiling was performed on archival formalin-fixed, paraffin-embedded (FFPE) kidney biopsy blocks stored in the Department of Pathology, Seoul National University Hospital, which were collected with the participants’ written informed consent. The study was approved by the Institutional Review Board of Seoul National University Hospital (IRB No. 2502-052-1611). Healthy donor controls were zero-time biopsies from 24 screened living kidney donors aged 36–66 years, obtained at transplantation; two sections from one control were combined into one profile. The DKD cohort comprised 28 biopsies from patients with diabetic kidney disease, with eGFR and urine protein-to-creatinine ratio (UPCR) at biopsy taken from linked hospital laboratory records. Twenty-four biopsies from patients with IgA nephropathy, profiled with the same panel, were used to train the PAS model described below.

### Spatial transcriptomic profiling

FFPE sections were profiled with Xenium In Situ (10x Genomics). Gene-specific padlock probes hybridize to target transcripts, ligated probes are amplified in place, and their barcodes are decoded through successive rounds of fluorescence imaging [13, 14]. Nuclear and multi-tissue staining guided cell segmentation, and decoded transcripts were assigned to segmented cells. The human 5K panel with a kidney-specific add-on provided 5,101 gene targets (Supplementary Methods). PAS-stained sections were digitized at 40× magnification (0.263 µm per pixel; Aperio GT450, Leica Biosystems).

Cells were analyzed in 11 annotated groups: collecting-duct groups CDPC, CDPCA, and CDPCB; distal convoluted tubule (DCT); endothelium (Endo); immune cells (IMM); parietal epithelial cells (PEC); podocytes (POD); proximal tubule (PT); stromal cells; and thick ascending limb (TAL), assigned in each specimen with reference to a single-nucleus RNA-sequencing dataset of 125 kidney biopsy samples from the same biobank (Supplementary Methods). For each biopsy, we recorded cell proportions, median cell area, transcripts captured per cell, cell-pair proximity, and cell-group pseudobulk expression. Proximity was the observed frequency of neighboring cell types relative to that expected from cell composition, summarized for four prespecified groups—immune–epithelial, endothelial–podocyte, stromal–epithelial, and epithelial–epithelial (Figure 1a)—and for each of 66 cell pairs. Proximity was available for 23 controls, and all other measurements for all 24 controls and 28 DKD biopsies.

**Figure 1.**
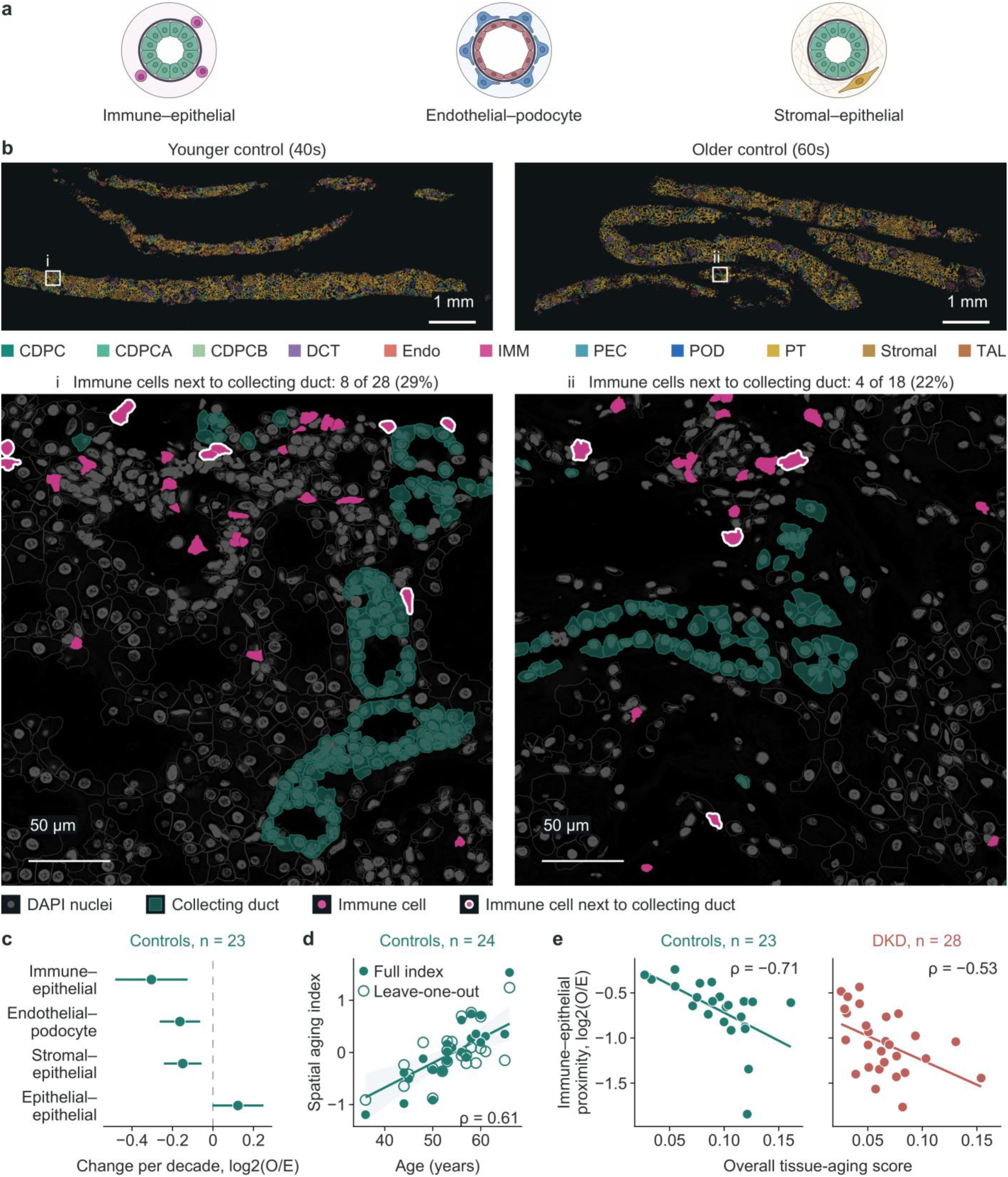
Spatial organization of healthy kidney aging. a: Schematic kidney microenvironments represented by the immune–epithelial, endothelial–podocyte, and stromal–epithelial proximity measurements. b: Cell-type maps of a younger (40s) and an older (60s) control biopsy, with cells colored by the 11 annotated groups, and a 300 × 300 µm region around collecting ducts from each, chosen among windows at least 80% covered by tissue as the window whose immune–collecting-duct contact was closest to the within-biopsy median. In the regions, registered DAPI nuclei are shown in dark gray, collecting-duct cells in green, and immune cells in magenta; immune cells with a collecting-duct neighbor are outlined in white, and their number and proportion among all immune cells in the region are shown above each region. The same linear DAPI display scale was used for both controls. Scale bars, 1 mm and 50 µm. c: Sex-adjusted change per decade in the four proximity groups among 23 controls; points are estimates and lines 95% confidence intervals. d: Spatial aging index against age in 24 controls (filled, full index; open, index built without that control) with the fitted line and 95% confidence band. e: Overall tissue-aging score against immune–epithelial proximity in controls (n = 23) and DKD (n = 28). Schematics in a were created with BioRender.com.

### Healthy control aging

Control-level proximity, cell proportions, cell area, transcript content, and pseudobulk expression were related to age per decade with sex adjustment; models for individual cell pairs also included the proportions of both participating cell groups. Upper-versus-lower age tertiles assessed direction, and false-discovery rates were controlled within measurement families [20]. A spatial aging index combined the control measurements most consistently associated with age; each control was also scored from an index built without that control.

### Control-derived tissue-aging scores

Within each cell group, the 20 genes that rose and the 20 that fell most consistently with control age (smallest sex-adjusted *P* values) were selected among genes detected in at least 15% of controls, after excluding signals attributable to a neighboring cell type. Each cell-group score was the mean log-transformed expression of the age-increased genes minus that of the age-decreased genes. The overall score averaged the 11 cell-group scores, and collecting-duct, tubular epithelial, tubulointerstitial, and filtration-barrier scores averaged their constituent groups (Supplementary Table S1). The scores contain no proximity or cell-area terms, and genes and weights were fixed in controls before any DKD outcome was examined.

### Estimation of tissue-aging scores from PAS histology

Each PAS-stained section was divided into 128 × 128 µm tiles, which were stain-normalized [21] and encoded with the UNI2-h pathology foundation model [18] (Figure 5a). Tile features were summarized for each biopsy, and a ridge regression model was trained to estimate the four compartment scores in 24 DKD and 24 IgA nephropathy biopsies, with each of these DKD biopsies scored by models built without that patient. Three DKD biopsies that were not used in training were scored by the model fitted to all training biopsies after alignment for a difference in PAS staining (Supplementary Methods).

### DKD analyses

For each DKD patient, the rate of eGFR change after biopsy (eGFR slope, in mL/min/1.73 m²per year) was estimated with the Theil–Sen method from measurements spanning at least 6 months. This was possible in 24 patients; the other 4 contributed to the tissue and baseline analyses. Measurements after kidney failure or transplantation were not used (Supplementary Methods). We used eGFR slope as the outcome because it reflects the pace of kidney function loss and is an accepted measure of chronic kidney disease progression [22, 23]. Each score was related to eGFR slope by partial Spearman correlation adjusted for age, sex, and baseline eGFR, with *P* values from 10,000 permutations [24], and in a second model additionally adjusted for log(1+UPCR). Sensitivity analyses are described in the Supplementary Methods. Associations of each score with eGFR and UPCR at biopsy were examined with adjustment for age and sex.

Within controls and within DKD, the overall score was correlated with each proximity group. To ask whether DKD tissue resembles older tissue, the DKD-versus-control difference in each of the 66 cell-pair proximities was compared with the age-related change of the same pair in controls, and the agreement was tested against permutations of control age.

PAS-estimated scores, available for all 24 patients in the slope analysis, were related to eGFR slope in the same way as the transcriptomic scores.

Expression changes were compared with control aging using genes outside all signatures, by relating control-age effects to DKD-versus-control effects and to overall-score effects within DKD (Supplementary Methods).

## Results

### Age-related changes in cell relationships and expression in control kidneys

Controls had a median age of 55 years (range, 36–66), and 10 were women. The 28 DKD patients had a median age of 50.5 years (range, 23–70), 8 were women, median baseline eGFR was 47.5 mL/min/1.73 m², median UPCR was 3.9 g/g, and median follow-up was 3.9 years. In the 24 patients with an eGFR slope, the median slope was −6.0 mL/min/1.73 m²per year (Table 1).

**Table 1.** Clinical characteristics and kidney-function follow-up in healthy donor controls and patients with DKD.

| **Characteristic** | **Healthy donor controls (n = 24)** | **DKD (n = 28)** |
|---|---|---|
| Age, years, median (range) | 55 (36–66) | 50.5 (23–70) |
| Women, n (%) | 10 (42) | 8 (29) |
| eGFR at biopsy, mL/min/1.73 m² | Normal (specific value not available) | 47.5 (36.6 to 73.8) |
| Peri-biopsy UPCR, g/g | Absent | 3.9 (3.0 to 7.7) |
| Follow-up from biopsy, years | – | 3.9 (1.3 to 6.4) |
| Post-biopsy eGFR dates used (n = 24) | – | 38 (15 to 48) |
| eGFR slope, mL/min/1.73 m²/year (n = 24) | – | −6.0 (−9.7 to −3.2) |
| Post-biopsy ACE inhibitor or ARB exposure | – | 26/28 |
| Post-biopsy SGLT2 inhibitor exposure | – | 7/28 |

Values are median (interquartile range) unless noted. The slope analysis required measurements spanning at least 6 months; all 28 DKD biopsies contributed to tissue and baseline analyses. eGFR measurements after kidney failure or transplantation were not used. One missing baseline UPCR was taken from day 10 after biopsy. Medication fields record any exposure after biopsy. ACE, angiotensin-converting enzyme; ARB, angiotensin receptor blocker; SGLT2, sodium-glucose cotransporter 2.

Among the 23 controls with proximity measurements, older age was associated with fewer contacts between immune and epithelial cells, between endothelial cells and podocytes, and between stromal and epithelial cells (sex-adjusted *q* = 0.0043 for each), whereas contacts among epithelial cells tended to increase (*q* = 0.055; Figure 1a,c; Supplementary Figure S1). In the tissue maps, immune cells lay next to collecting-duct cells less often in the older control (Figure 1b). Immune–collecting-duct and PT–TAL contacts declined even after accounting for the abundance of each cell group (Supplementary Figure S2), and cell proportions changed little with age (Supplementary Figure S1). Aging therefore changed where cells lie relative to one another more than how many cells of each type were present. A spatial aging index combining these measurements rose with age (0.72 units per decade; 95% CI, 0.51–0.92), also when each control was scored from an index built without that control (ρ = 0.61; Figure 1d).

All five tissue-aging scores rose across the control age range (overall score, 1.14 SD per decade; Figure 2c; Supplementary Table S1). Several age-increased genes recurred across cell groups: *FOXO3* rose with age in proximal tubule, thick ascending limb, endothelium, parietal epithelium, and stromal cells, and *IRAK1* in tubular, parietal epithelial, and stromal cells. The cell-cycle inhibitor *CDKN1A* rose in proximal tubule, the pro-apoptotic *BBC3* in thick ascending limb, the mitophagy regulator *PINK1* in endothelium and podocytes, and *SOCS1*, *MDM2*, and *DDB2* in immune cells (Figure 2a). In proximal tubule, *FOXO3* and *CDKN1A* transcripts per cell were higher in the older than in the younger control (Figure 2b). Stromal cells gained immune-lineage and matrix-remodeling transcripts (*CXCR4*, *RUNX3*, *IKZF1*, *MMP14*, *HTRA1*), and endothelial cells gained interferon-inducible transcripts (*CYBB*, *SAMHD1*). Age-associated pathways differed by cell group, with oxidative phosphorylation and fatty-acid metabolism in proximal tubule and collecting duct, and interferon and adaptive-immune programs in immune, endothelial, and stromal cells; the SenMayo senescence signature increased with age only in podocytes (Supplementary Figure S3). Proximal-tubule cells resembling the injured PT state of the single-nucleus reference, with lower transport-gene and higher stress-marker expression, also became more frequent with age (0.040 per decade; 95% CI, 0.015–0.064); in DKD, a higher burden of these cells was associated with lower eGFR at biopsy but not with the subsequent eGFR slope (Supplementary Figure S4).

**Figure 2.**
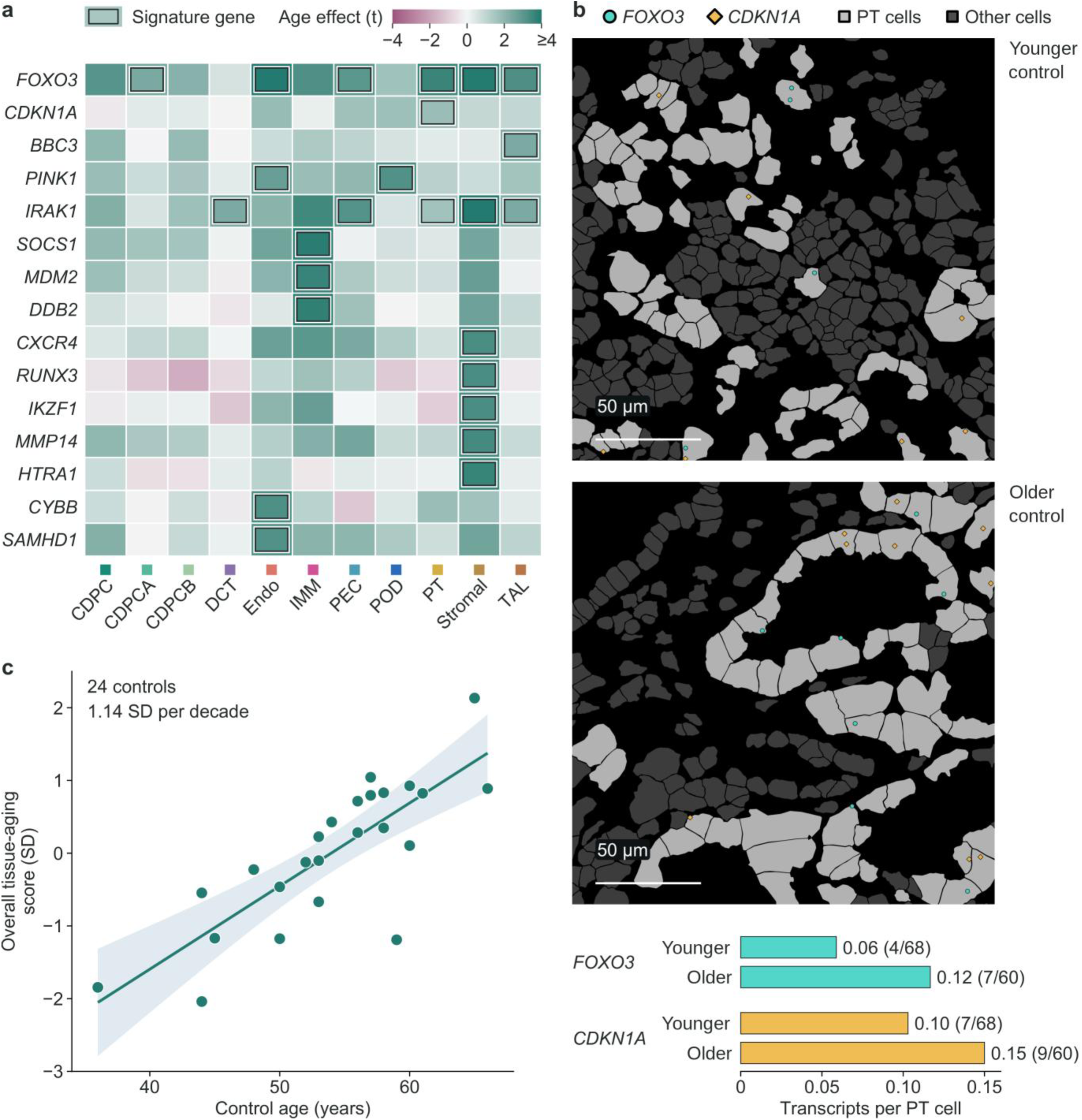
Cell-group expression programs of healthy kidney aging. a: Sex-adjusted age effects (t statistics) for selected age-increased genes across the 11 cell groups in 24 controls; outlined cells mark genes included in that cell group’s fixed signature. b: *FOXO3* (turquoise circles) and *CDKN1A* (gold diamonds) transcripts over proximal-tubule regions (proximal-tubule cells in light gray, other cells in dark gray) of the younger and older control shown in Figure 1b, selected as the window closest to the within-biopsy median of these transcripts per PT cell; only transcripts assigned to PT cells with centroids in the window are shown, and those PT cells are light gray. Bars give the displayed transcript counts divided by the number of those PT cells, with transcript counts/PT-cell counts in parentheses. Overlapping transcript symbols are separated by at most 2 µm for display. Scale bars, 50 µm. c: Overall tissue-aging score against age in 24 controls with the fitted line and 95% confidence band.

### Spatial and expression changes in DKD

Changes in cell contacts in DKD largely followed the direction of control aging. Cell pairs whose contact decreased with age in controls also tended to be less in contact in DKD than in controls, and pairs whose contact increased with age tended to be more in contact (ρ = 0.65 across the 66 cell pairs; 80.3% of pairs in the same direction; permutation *P* = 0.0066; Figure 3a). Immune–collecting-duct, immune–PT, endothelial–podocyte, and parietal epithelial–stromal contacts were lower both in older controls and in DKD (Figure 3b).

**Figure 3.**
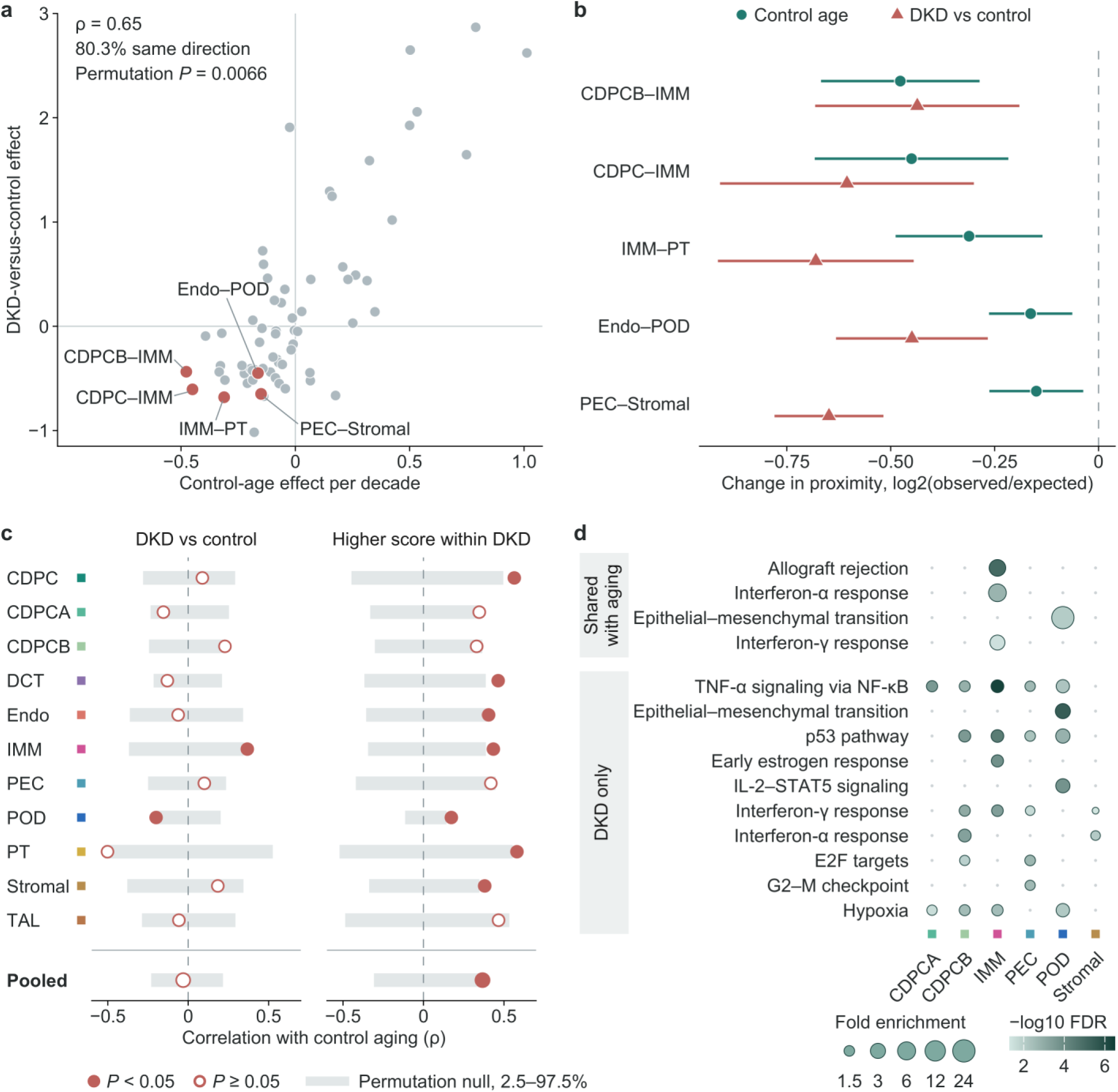
Spatial and expression changes in DKD compared with control aging. a: Sex-adjusted control-age effect against the age- and sex-adjusted DKD-versus-control effect for 66 cell-pair proximity measurements (ρ = 0.65; 80.3% of pairs in the same direction; permutation *P* = 0.0066); labeled pairs are shown in b. b: Control-age and DKD-versus-control effects with 95% confidence intervals for five anatomical pairs. c: Correlation between control-age effects and DKD-versus-control effects (left) or overall-score effects within DKD (right) across genes outside the signatures, by cell group and pooled. Gray bars show the 2.5th–97.5th percentiles of 1,000 permutations; filled points, permutation *P* < 0.05. d: Hallmark pathways enriched among genes that differed between DKD and control tissue, separated into those that changed in the same direction with control age and those that did not. Point size shows fold enrichment and color shows false discovery rate.

Expression changes in DKD were less aligned with control aging than the spatial changes. Outside the signature genes, differences between DKD and control tissue showed no overall correspondence with control-age effects (ρ = −0.03; Figure 3c), except in immune cells (ρ = 0.37), where the shared changes involved interferon and T-cell activation programs. Changes specific to DKD involved TNF-α–NF-κB, hypoxia, p53, and cell-cycle programs in collecting-duct, parietal epithelial, podocyte, and immune cells (Figure 3d). Within DKD, biopsies with a higher overall score showed aging-direction changes in genes outside the signature in all 11 cell groups (ρ = 0.37, permutation *P* = 0.008; Figure 3c).

Biopsies with a higher overall tissue-aging score had fewer immune–epithelial contacts, both in controls (ρ = −0.71, n = 23) and in DKD (ρ = −0.53, n = 28; Figure 1e), so that the expression-based score and the spatial changes pointed in the same direction. In DKD, the filtration-barrier, endothelial, immune, podocyte, and stromal scores were associated with lower eGFR at biopsy, but the overall score showed no clear association with patient age, eGFR, or proteinuria at biopsy (standardized β = 0.23, −0.25, and 0.18; Supplementary Table S2). The overall score therefore reflected a property of the tissue that these clinical measures did not capture.

### Tissue-aging scores and eGFR decline in DKD

After adjustment for age, sex, and baseline eGFR, patients with a higher overall tissue-aging score lost eGFR faster (partial ρ = −0.63; permutation *P* = 0.002; Figure 4a,b). All four compartment scores showed this association (partial ρ, −0.58 to −0.69), as did the scores of every cell group except podocytes (−0.49 to −0.72, versus −0.10 for podocytes; Figure 4a; Supplementary Table S3). Results were similar in sensitivity analyses, and additional adjustment for proteinuria did not change the association (overall score, partial ρ = −0.61; permutation *P* = 0.0045).

**Figure 4.**
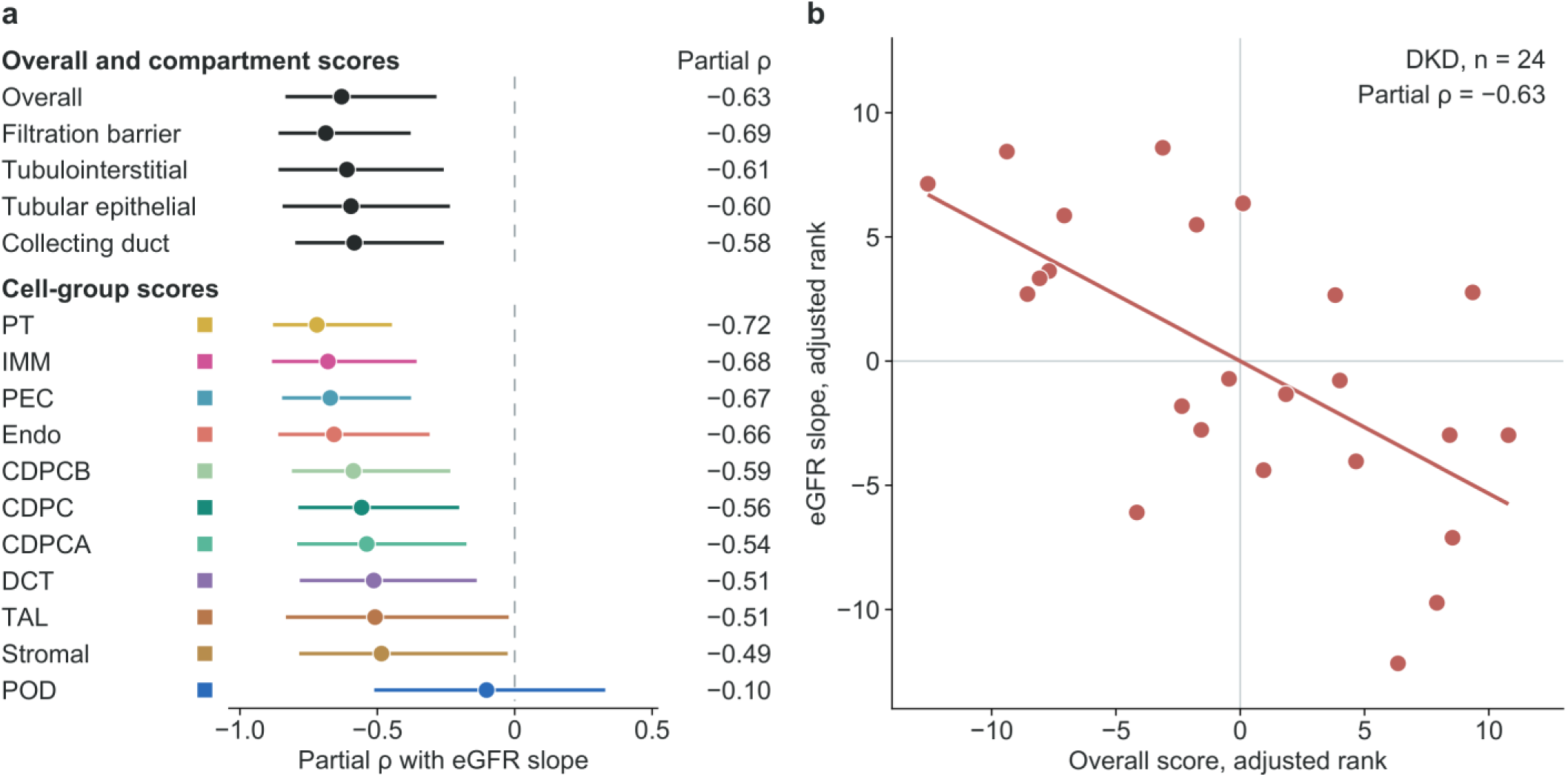
Tissue-aging scores and eGFR decline in DKD. a: Partial Spearman correlation of each of the 16 tissue-aging scores with eGFR slope in 24 patients, adjusted for age, sex, and baseline eGFR; lines show patient-bootstrap 95% confidence intervals from 2,000 resamples, with ranking and covariate adjustment repeated in each resample; cell-group points are colored as in Figure 1. b: Ranks of the overall score and eGFR slope after adjustment for the same covariates.

### Tissue-aging scores estimated from PAS histology

In the 24 DKD biopsies used for training, scores estimated from PAS slides correlated with the transcriptomic compartment scores (mean Spearman ρ = 0.62; highest for the filtration barrier, 0.72, and lowest for the collecting duct, 0.59; Figure 5b; Supplementary Figure S5). In the 24 patients with an eGFR slope, higher PAS-estimated scores were associated with faster eGFR loss for all four compartments (partial ρ, −0.44 to −0.56; permutation *P* = 0.010–0.043), close to the associations of the transcriptomic scores (−0.58 to −0.69; Figure 5c; Supplementary Table S4). Additional adjustment for proteinuria attenuated these associations only slightly, and they remained significant for the tubulointerstitial, tubular epithelial, and filtration-barrier scores (partial ρ, −0.44 to −0.49; collecting duct, −0.37; Supplementary Table S4). Tiles with the highest estimates showed cortical tubules with thickened, wrinkled basement membranes and a widened interstitium, and these tiles were largely the same for the filtration-barrier and tubulointerstitial estimates (Supplementary Figure S6f,g). Blockwise occlusion localized higher-estimate contributions to PAS-positive tubular basement membranes and intervening interstitium, and lower-estimate contributions to tubular lumens and epithelium across control, DKD, and IgA nephropathy examples (Figure 6).

**Figure 5.**
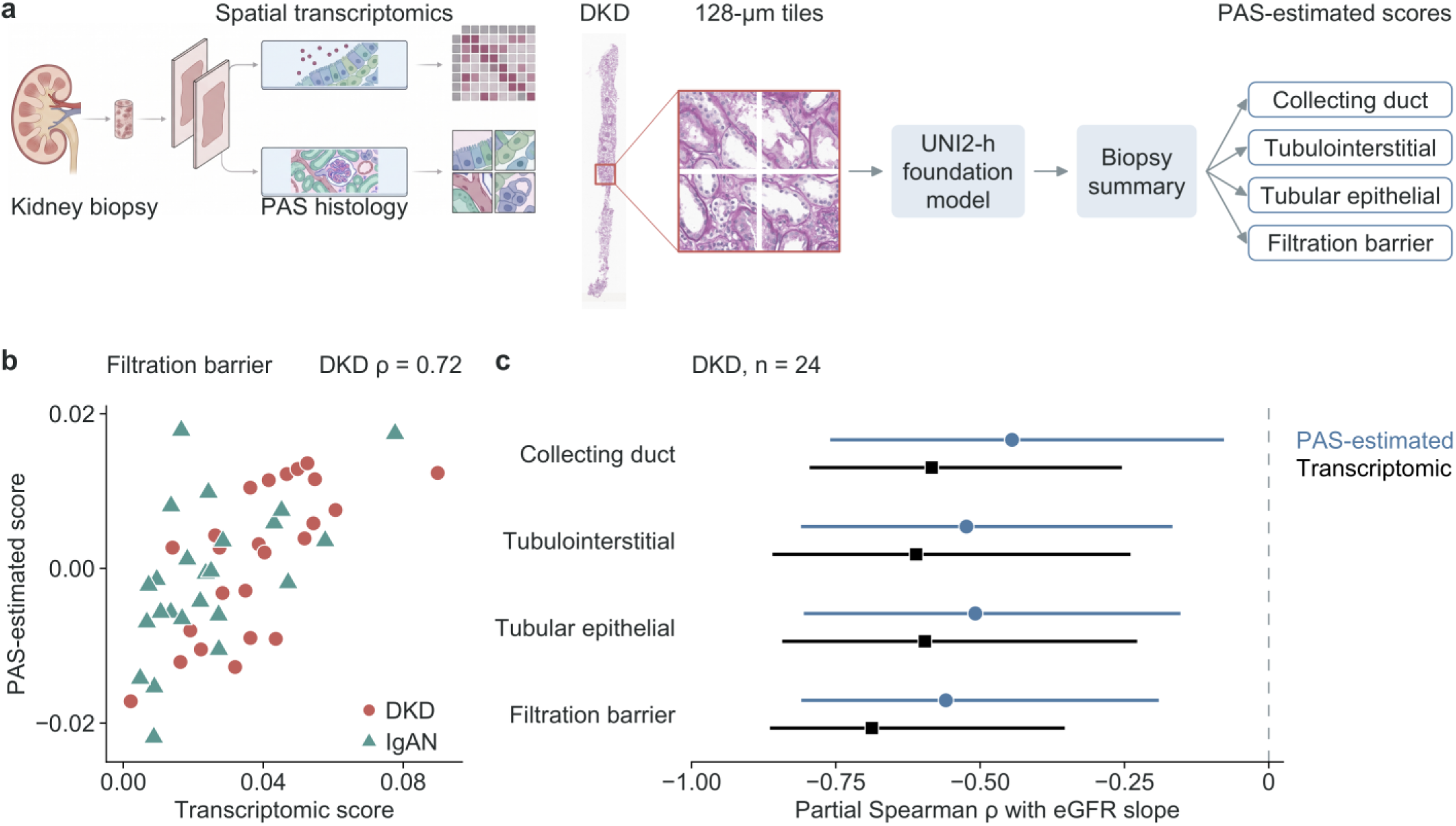
Estimation of tissue-aging scores from PAS histology. a: Schematic of spatial transcriptomic measurement and PAS-based estimation, followed by the analyzed PAS section and image tiles. The red box marks the location of the displayed tiles. PAS slides were divided into 128-µm tiles, encoded with a pathology foundation model, summarized per biopsy, and mapped to the four compartment scores; models were trained on 48 biopsies (24 DKD, 24 IgA nephropathy), and each of these DKD biopsies was scored by models built without that patient; three additional DKD biopsies were scored by the model fitted to all 48 biopsies. b: PAS-estimated against transcriptomic filtration-barrier score in DKD (red) and IgA nephropathy (jade); scores were centered within each disease during training. c: Partial Spearman correlations of the four PAS-estimated scores (steel-blue circles) and their transcriptomic counterparts (black squares) with eGFR slope in the same 24 DKD patients, adjusted for age, sex, and baseline eGFR. Lines show percentile 95% confidence intervals from 2,000 patient-level bootstrap resamples, with ranking and adjustment repeated in each resample. Schematics in a were created with BioRender.com.

**Figure 6.**
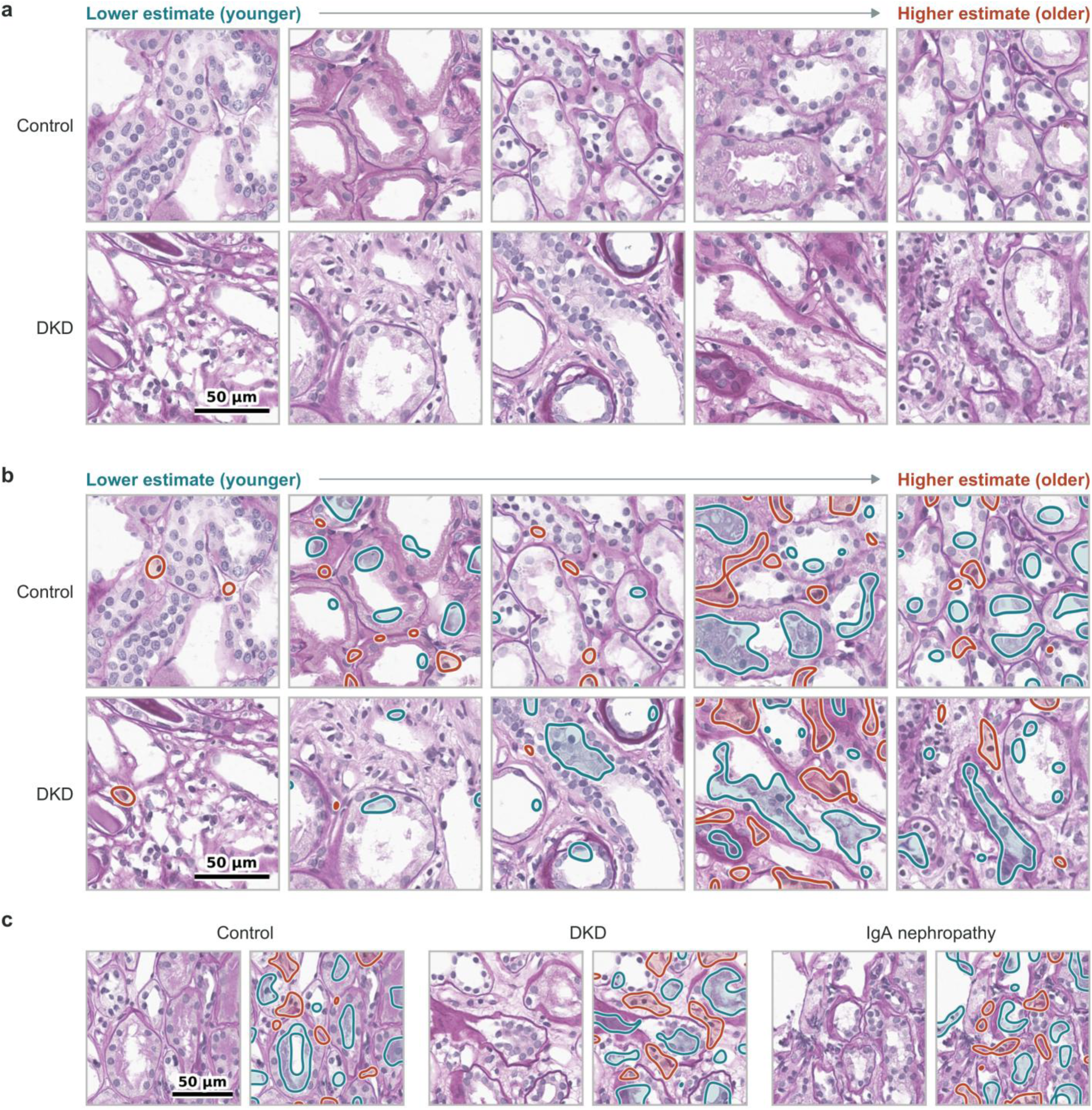
What the PAS model reads as younger and older tissue. a: PAS tiles ordered by the tile-level tissue-aging estimate from the lowest to the highest quintile, in controls and DKD. b: The same tiles with occlusion outlines; each 16-µm block was flattened and the tile re-embedded. Red marks regions whose removal lowers the estimate (older-making), and blue marks regions whose removal raises it (younger-making). c: The highest-estimate tile in control, DKD, and IgA nephropathy without and with outlines. Older-making regions fall on PAS-positive tubular basement membranes and the interstitium between tubules; younger-making regions fall within tubular lumens and epithelium. Scale bars, 50 µm.

## Discussion

Healthy donor kidneys showed selective age-related changes in the arrangement of cells, together with a coordinated stress-response expression program in the same cell groups. DKD tissue shared this spatial pattern, and DKD kidneys with a stronger aging phenotype lost function faster. The phenotype could be estimated in part from the routine PAS slide.

Kidney aging is usually described by lesions such as glomerulosclerosis and interstitial fibrosis [1]. The changes found here are at a finer scale: cells change their neighbors and expression programs while their numbers are preserved. In DKD, this aging phenotype was largely separate from chronological age, eGFR, and proteinuria, suggesting that patients with similar clinical measures can have kidneys that differ in biological age and in their capacity to withstand diabetic injury. That the tubular, endothelial, immune, and parietal epithelial scores, but not the podocyte score, carried this information points to the tubulointerstitium and the glomerular microenvironment, rather than podocyte loss alone, as the site of this vulnerability.

The age-increased genes include *FOXO3*, *CDKN1A*, *BBC3*, *PINK1*, *IRAK1*, and *SOCS1*, consistent with stress-response and cell-cycle regulation in the aging kidney [5, 6, 8]. *FOXO3* and *CDKN1A* (p21) mediate growth arrest, *PINK1* mitochondrial quality control, and *IRAK1* and *SOCS1* innate-immune signaling; their rise across tubular, endothelial, and stromal cells suggests a stress response shared across the kidney, whereas the SenMayo senescence signature rose only in podocytes. Because the signature was defined by age in kidneys without established disease, it provides a reference for healthy aging rather than for injury. Unlike the injured PT state, which related to eGFR at biopsy rather than to later decline, the aging score related to later decline, so it is unlikely to be simply a measure of current tubular injury. With genes selected by rank, the signature reflects a coordinated program rather than individually validated genes. Inflammation, tubular stress, and interstitial remodeling contribute to DKD progression [9, 10, 12]. Aging and DKD therefore appear to converge on similar tissue arrangements through largely different expression programs, with interferon and T-cell activation in immune cells as the main shared component; within DKD, the aging axis extended beyond the signature genes, so the score marks a broader state of the tissue rather than a small set of genes.

Kidney disease in people with diabetes is usually attributed to diabetes without a biopsy when another cause is not evident, and the role of biopsy and biomarkers in refining this diagnosis remains under investigation [25]. Histologic classification describes the glomerular lesions of diabetic nephropathy alongside interstitial and vascular injury [26] but does not grade the age-related state of the tissue. Estimating this state from the PAS slide already obtained for diagnosis would make it usable in routine practice. The tiles that drove the highest estimates showed tubular basement membrane thickening and interstitial widening (Figure 6), features of chronic tubulointerstitial change, and were similar across compartments. The PAS model therefore appears to read a shared tubulointerstitial appearance that accompanies the molecular aging program, rather than compartment-specific lesions, and because it was trained on molecular scores, it may capture gradations of this appearance that are not graded on routine review. This may be relevant to current DKD care. With SGLT2 inhibitors, nonsteroidal mineralocorticoid receptor antagonists, and GLP-1 receptor agonists now available for DKD [25], selecting patients for earlier or combined therapy depends on identifying those likely to progress, and the associations observed here support evaluating PAS-estimated tissue-aging scores together with clinical measures. Testing this requires prospective cohorts in which the PAS-estimated score is validated against tissue measurements and compared with clinical and histologic predictors.

This study has several strengths. Zero-time biopsies from 24 screened healthy donor controls provide a reference for aging without established kidney disease across three decades of adult life. Although donor tissue obtained at surgery shows higher expression of immediate-early stress genes than percutaneous biopsies [27], the signature was defined by age among donors who underwent the same procedure. The 5,101-gene spatial panel measures cellular identity, expression, and neighbors in the same section, so molecular scores and spatial organization could be compared within each biopsy. The DKD cohort came from the same biobank with linked laboratory follow-up, the gene sets and weights were fixed in controls before any DKD outcome was examined, and each PAS-estimated score came from a model that excluded that patient.

This study has several limitations. Controls spanned 36–66 years, and gene selection and control-age assessment used the same controls. The slope and PAS analyses included 24 patients, and PAS-estimated scores have not been tested in an external cohort. Scores correlated with transcripts captured per cell in DKD, which may reflect assay efficiency, cellular RNA content, or tissue composition. Diabetes duration, glycemic control, blood pressure, and treatment at biopsy were unavailable; the cohort represents biopsied DKD with substantial proteinuria and reduced eGFR; and proximity depends on segmentation and cell annotation.

A tissue-aging score obtained from the diagnostic biopsy, and in part from its PAS slide, could help identify patients with DKD whose kidneys are biologically older than their clinical measures suggest and who may benefit from earlier or intensified kidney-protective therapy.

## Data availability

Single-nucleus RNA sequencing data used as the reference will be deposited in the Korea BioData Station (K-BDS) with controlled access; access will be granted upon application and approval. Individual-level spatial transcriptomic, PAS image and clinical data are not publicly available. The archival biopsy tissue was stored and used for research based on the patients’ written consent for donation of human biospecimens, and neither this consent nor the approval of the Institutional Review Board of Seoul National University Hospital covers public release or transfer outside Korea of individual-level data generated from the tissue. De-identified data can be shared through a collaboration agreement with the KORNERSTONE investigators, subject to approval by the Institutional Review Board and the Biobank of Seoul National University Hospital; requests should be addressed to the corresponding authors. Gene-level and cell type-level summary statistics are provided in the Supplementary Tables.

## Supplementary information

**Supplementary Table S1.** Definition and control-age associations of the tissue-aging scores applied to DKD (with fixed signature genes by cell group)

| **Tissue-aging score** | **Constituent cell groups** | **n** | **Age β (95% CI)** | ***P*** |
|---|---|---|---|---|
| Overall tissue-aging score | CDPC, CDPCA, CDPCB, DCT, Endo, IMM, PEC, POD, PT, Stromal, TAL | 24 | 1.14 (0.76 to 1.53) | <0.001 |
| Collecting-duct score | CDPC, CDPCA, CDPCB | 24 | 1.12 (0.72 to 1.52) | <0.001 |
| Tubulointerstitial score | PT, TAL, DCT, Stromal | 24 | 1.05 (0.61 to 1.48) | <0.001 |
| Tubular epithelial score | DCT, PT, TAL | 24 | 0.94 (0.46 to 1.42) | <0.001 |
| Filtration-barrier score | Endo, POD, PEC | 24 | 1.18 (0.82 to 1.54) | <0.001 |
| **Cell group** | **Age-increased genes** | **Age-decreased genes** |  |  |
| CDPC (20/20) | *ERP27*, *PRKACB*, *SLFN5*, *UCHL1*, *TOX*, *UCP2*, *SHROOM1*, *ST6GAL1*, *STX7*, *PKIA*, *ALDH3A2*, *KIF2A*, *CHEK1*, *JAK1*, *MVP*, *C1RL*, *FLOT2*, *CTTNBP2NL*, *NUAK2*, *CASP10* | *PAX5*, *FOXN1*, *MUC5B*, *RELT*, *TBX15*, *KLK10*, *TNFRSF9*, *TAC4*, *ADM*, *SSX4*, *FCRL4*, *PCDH8*, *UNC5A*, *RORB*, *GPR55*, *PAX7*, *DNAH8*, *PCSK9*, *TBX21*, *ZNF165* |  |  |
| CDPCA (20/20) | *CALB1*, *PDCD4*, *MAOA*, *GOLPH3*, *HOXB7*, *PRXL2A*, *FAM107A*, *NDUFAB1*, *STK16*, *MPZL1*, *BAALC*, *VLDLR*, *TSPAN3*, *BARX2*, *FAM131C*, *FOXO3*, *MTCH2*, *RWDD1*, *HPRT1*, *AKTIP* | *CD22*, *MRAP*, *ADAMTS5*, *CMTM5*, *ZNF165*, *NCR3*, *PRG4*, *CD1A*, *HAMP*, *DAPK2*, *VIPR2*, *EFS*, *CNTN2*, *ITIH1*, *TRPC5*, *OPALIN*, *DDX25*, *LRRC3*, *CALB2*, *CHRNB4* |  |  |
| CDPCB (20/20) | *UCHL1*, *ALDH3A2*, *CEP19*, *RAI2*, *CALB1*, *PRKAG1*, *PRKACB*, *MCCC1*, *EIF5A*, *NPM3*, *TUBB*, *CYFIP2*, *HOXB7*, *VLDLR*, *ADAMTSL3*, *THAP11*, *EMC4*, *PDHA1*, *SAT2*, *VEGFB* | *PTPN5*, *DEFB103B*, *SH3TC2*, *PLK1*, *SCGB1D2*, *CST5*, *CHST8*, *MUC5B*, *CDSN*, *IL25*, *PAGE4*, *CBLN4*, *ERAS*, *LRTM1*, *RS1*, *HTR1B*, *FASLG*, *ZSCAN1*, *GPR149*, *NELL1* |  |  |
| DCT (20/20) | *CALB1*, *SEPTIN9*, *IRAK1*, *HOXB7*, *F2*, *BMP8B*, *CD99*, *CBR1*, *KCNE1*, *EIF5A*, *NRSN2*, *TSPAN3*, *LAPTM4B*, *KLRF1*, *HPRT1*, *STUB1*, *EID1*, *THAP11*, *HMGB2*, *STMN3* | *CALR3*, *KLK15*, *IL17F*, *CCL23*, *BCAR4*, *P2RX3*, *INSL4*, *KCNH4*, *IL24*, *ACOD1*, *AQP8*, *SPIB*, *PRM1*, *CTCFL*, *ITPKA*, *DAB1*, *HEPACAM*, *CYP1A2*, *PAEP*, *MC3R* |  |  |
| Endo (20/20) | *FOXO3*, *NDRG4*, *INMT*, *CYBB*, *SAMHD1*, *FAM107A*, *CCND3*, *LDHA*, *PMP22*, *ABCG2*, *CD302*, *EVI2A*, *EVI2B*, *NGFR*, *PINK1*, *STEAP4*, *PNPLA2*, *SRPX*, *CD59*, *MT1X* | *NETO2*, *GLP1R*, *WNT1*, *CRYBB3*, *DLK2*, *PDE4B*, *IL21R*, *PLEKHO1*, *VIPR2*, *CACNG5*, *CHODL*, *RASSF2*, *CNNM1*, *SLC5A7*, *SSX4*, *SMOX*, *TSPO*, *ALOX12*, *ATP2B1*, *APLNR* |  |  |
| IMM (20/20) | *SLC25A40*, *AHR*, *UBD*, *SNX30*, *PSTPIP2*, *PPP1R7*, *RUVBL2*, *SOCS1*, *DDB2*, *ALCAM*, *TSC2*, *RNF167*, *MDM2*, *SMAP2*, *NOP2*, *RRS1*, *PIP4K2A*, *INO80D*, *SMARCA2*, *CDC73* | *GUCY1A2*, *CD248*, *SEPTIN5*, *SYP*, *WFDC1*, *CCN4*, *PADI2*, *RILPL1*, *LANCL2*, *DLL4*, *MCAM*, *ERAS*, *MALL*, *CACNA1C*, *MRTFB*, *AR*, *CDK3*, *SSX2*, *PALMD*, *EDNRA* |  |  |
| **Cell group** | **Age-increased genes** | **Age-decreased genes** |  |  |
| PEC (20/20) | *FLOT2*, *DUSP23*, *EPDR1*, *AP2M1*, *TNMD*, *EIF5A*, *MAOA*, *TGFBR1*, *PROS1*, *C1RL*, *IRAK1*, *SERPINA3*, *NRXN2*, *FAIM*, *SOD3*, *GCLM*, *FOXO3*, *ALDOA*, *MTCH2*, *HOXB7* | *SPRY2*, *CLEC4M*, *BMP7*, *DHRS2*, *ADAM33*, *DRC1*, *WT1*, *ABCG1*, *NR4A3*, *RGS4*, *SLIT3*, *CRB2*, *MYLK2*, *CAMK2B*, *ZNF469*, *SYCP1*, *COL11A1*, *F5*, *TNNT2*, *CD83* |  |  |
| POD (20/20) | *FAM107A*, *S1PR1*, *PRDM1*, *CLIC4*, *RFTN1*, *EPAS1*, *PINK1*, *IL3RA*, *PIP4K2A*, *CAVIN2*, *NT5E*, *C1QTNF1*, *AHR*, *GAS2L1*, *FGR*, *GIMAP4*, *MYLK3*, *PECAM1*, *MCM3*, *CD109* | *SCGB1D2*, *CALB2*, *TRIB3*, *HCN4*, *LRTM1*, *TNNI3*, *CACNG4*, *GPR26*, *PDLIM2*, *MUC16*, *BAD*, *GADD45A*, *CDX1*, *VSTM1*, *HDAC5*, *TRPM8*, *KLK10*, *PAX1*, *RHEB*, *CCR8* |  |  |
| PT (20/20) | *FOXO3*, *LBH*, *HOXB7*, *KCNC3*, *TBC1D8B*, *CDKN1A*, *PRXL2A*, *MAOA*, *APELA*, *PRNP*, *IRAK1*, *FLOT2*, *EEF1G*, *FOXO4*, *DAP*, *CNP*, *VSIR*, *TMEM100*, *MCM5*, *SSBP3* | *STRIP2*, *SSTR2*, *CD83*, *NPC1L1*, *CCR1*, *CPEB1*, *SCN7A*, *BDKRB1*, *MYOT*, *TH*, *CALR3*, *ZC3H12D*, *IL10*, *CCL23*, *MAML2*, *BNC2*, *SLC19A1*, *GRID1*, *PHETA1*, *KRT13* |  |  |
| Stromal (20/20) | *CCND3*, *IRAK1*, *FOXO3*, *CFB*, *INMT*, *HTRA1*, *TPBG*, *IRS2*, *HOPX*, *C1RL*, *MMP14*, *CYP1B1*, *STEAP4*, *SEPTIN9*, *CXCR4*, *RUNX3*, *RRS1*, *IKZF1*, *SBNO2*, *DPYSL2* | *GUCY1A2*, *EEF1A2*, *STC2*, *EGFL7*, *YIF1B*, *NR4A3*, *NPY1R*, *SLC34A1*, *NKX3*-1, *SLC13A3*, *SLC22A8*, *PRKCG*, *LRP2*, *PDGFB*, *KCNQ2*, *FLT4*, *PAEP*, *CYP27B1*, *BMP8A*, *CD44* |  |  |
| TAL (20/20) | *FOXO3*, *ABI3*, *PPP1R1B*, *NCS1*, *CCND3*, *ZFYVE21*, *TGFA*, *STAT5A*, *HOXB7*, *SMAD7*, *PRXL2A*, *BLOC1S2*, *BBC3*, *TSPAN8*, *IRAK1*, *NDRG2*, *TRAF3*, *MAOA*, *CNP*, *SEPTIN9* | *JAG1*, *VSTM1*, *FGF4*, *TEKT1*, *SAA4*, *RFPL3*, *KCNH4*, *SERPINC1*, *SAPCD2*, *HTR3A*, *NPSR1*, *SFRP4*, *NLRP5*, *ECEL1*, *TRPC7*, *PCK1*, *FOXE1*, *CD83*, *MECOM*, *BRCA1* |  |  |

Scores are unweighted means of their constituent cell-group scores; control age coefficients are SD per decade, adjusted for sex. A score from the age-increased genes alone correlated with the full score at r=0.983 across 52 biopsies (controls, 0.975; DKD, 0.996). Many age-decreased genes are expressed at low levels.
20 genes per direction were ranked by sex-adjusted control-age *P* within cell group, after detection and neighboring-cell-group specificity checks. Counts in parentheses are age-increased/age-decreased genes; selection by rank did not impose a gene-level FDR cutoff.
20 genes per direction were ranked by sex-adjusted control-age *P* within cell group, after detection and neighboring-cell-group specificity checks. Counts in parentheses are age-increased/age-decreased genes; selection by rank did not impose a gene-level FDR cutoff.

**Supplementary Table S2.**
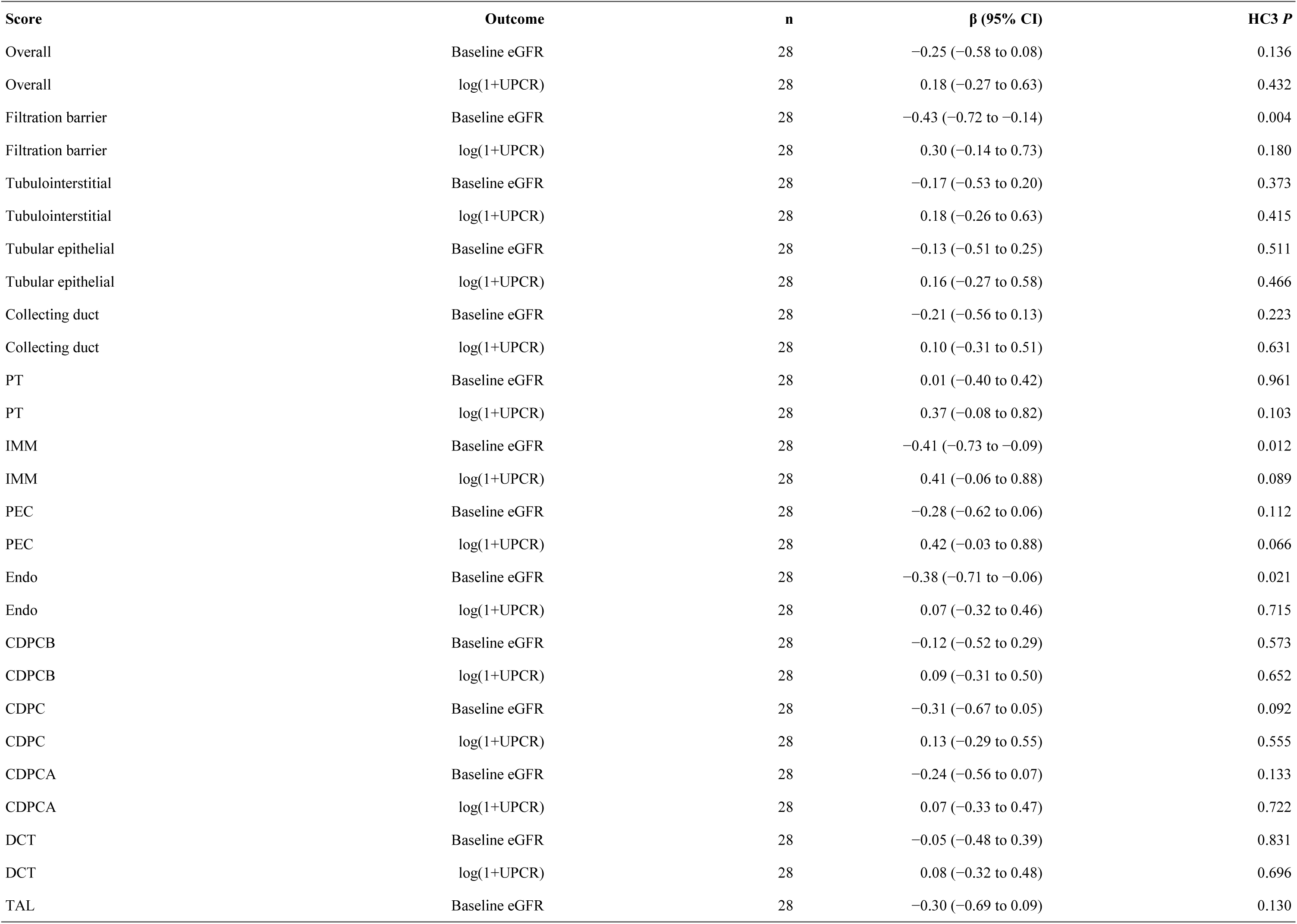
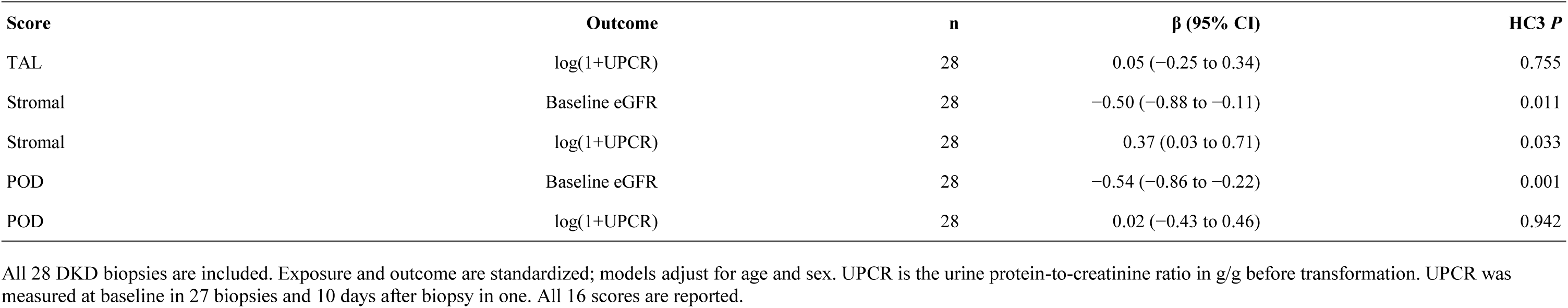
Tissue-aging scores and baseline eGFR and proteinuria in DKD.

| **Score** | **Outcome** | **n** | **β (95% CI)** | **HC3 *P*** |
|---|---|---|---|---|
| Overall | Baseline eGFR | 28 | −0.25 (−0.58 to 0.08) | 0.136 |
| Overall | log(1+UPCR) | 28 | 0.18 (−0.27 to 0.63) | 0.432 |
| Filtration barrier | Baseline eGFR | 28 | −0.43 (−0.72 to −0.14) | 0.004 |
| Filtration barrier | log(1+UPCR) | 28 | 0.30 (−0.14 to 0.73) | 0.180 |
| Tubulointerstitial | Baseline eGFR | 28 | −0.17 (−0.53 to 0.20) | 0.373 |
| Tubulointerstitial | log(1+UPCR) | 28 | 0.18 (−0.26 to 0.63) | 0.415 |
| Tubular epithelial | Baseline eGFR | 28 | −0.13 (−0.51 to 0.25) | 0.511 |
| Tubular epithelial | log(1+UPCR) | 28 | 0.16 (−0.27 to 0.58) | 0.466 |
| Collecting duct | Baseline eGFR | 28 | −0.21 (−0.56 to 0.13) | 0.223 |
| Collecting duct | log(1+UPCR) | 28 | 0.10 (−0.31 to 0.51) | 0.631 |
| PT | Baseline eGFR | 28 | 0.01 (−0.40 to 0.42) | 0.961 |
| PT | log(1+UPCR) | 28 | 0.37 (−0.08 to 0.82) | 0.103 |
| IMM | Baseline eGFR | 28 | −0.41 (−0.73 to −0.09) | 0.012 |
| IMM | log(1+UPCR) | 28 | 0.41 (−0.06 to 0.88) | 0.089 |
| PEC | Baseline eGFR | 28 | −0.28 (−0.62 to 0.06) | 0.112 |
| PEC | log(1+UPCR) | 28 | 0.42 (−0.03 to 0.88) | 0.066 |
| Endo | Baseline eGFR | 28 | −0.38 (−0.71 to −0.06) | 0.021 |
| Endo | log(1+UPCR) | 28 | 0.07 (−0.32 to 0.46) | 0.715 |
| CDPCB | Baseline eGFR | 28 | −0.12 (−0.52 to 0.29) | 0.573 |
| CDPCB | log(1+UPCR) | 28 | 0.09 (−0.31 to 0.50) | 0.652 |
| CDPC | Baseline eGFR | 28 | −0.31 (−0.67 to 0.05) | 0.092 |
| CDPC | log(1+UPCR) | 28 | 0.13 (−0.29 to 0.55) | 0.555 |
| CDPCA | Baseline eGFR | 28 | −0.24 (−0.56 to 0.07) | 0.133 |
| CDPCA | log(1+UPCR) | 28 | 0.07 (−0.33 to 0.47) | 0.722 |
| DCT | Baseline eGFR | 28 | −0.05 (−0.48 to 0.39) | 0.831 |
| DCT | log(1+UPCR) | 28 | 0.08 (−0.32 to 0.48) | 0.696 |
| TAL | Baseline eGFR | 28 | −0.30 (−0.69 to 0.09) | 0.130 |
| TAL | log(1+UPCR) | 28 | 0.05 (−0.25 to 0.34) | 0.755 |
| Stromal | Baseline eGFR | 28 | −0.50 (−0.88 to −0.11) | 0.011 |
| Stromal | log(1+UPCR) | 28 | 0.37 (0.03 to 0.71) | 0.033 |
| POD | Baseline eGFR | 28 | −0.54 (−0.86 to −0.22) | 0.001 |
| POD | log(1+UPCR) | 28 | 0.02 (−0.43 to 0.46) | 0.942 |

All 28 DKD biopsies are included. Exposure and outcome are standardized; models adjust for age and sex. UPCR is the urine protein-to-creatinine ratio in g/g before transformation. UPCR was measured at baseline in 27 biopsies and 10 days after biopsy in one. All 16 scores are reported.

**Supplementary Table S3.**
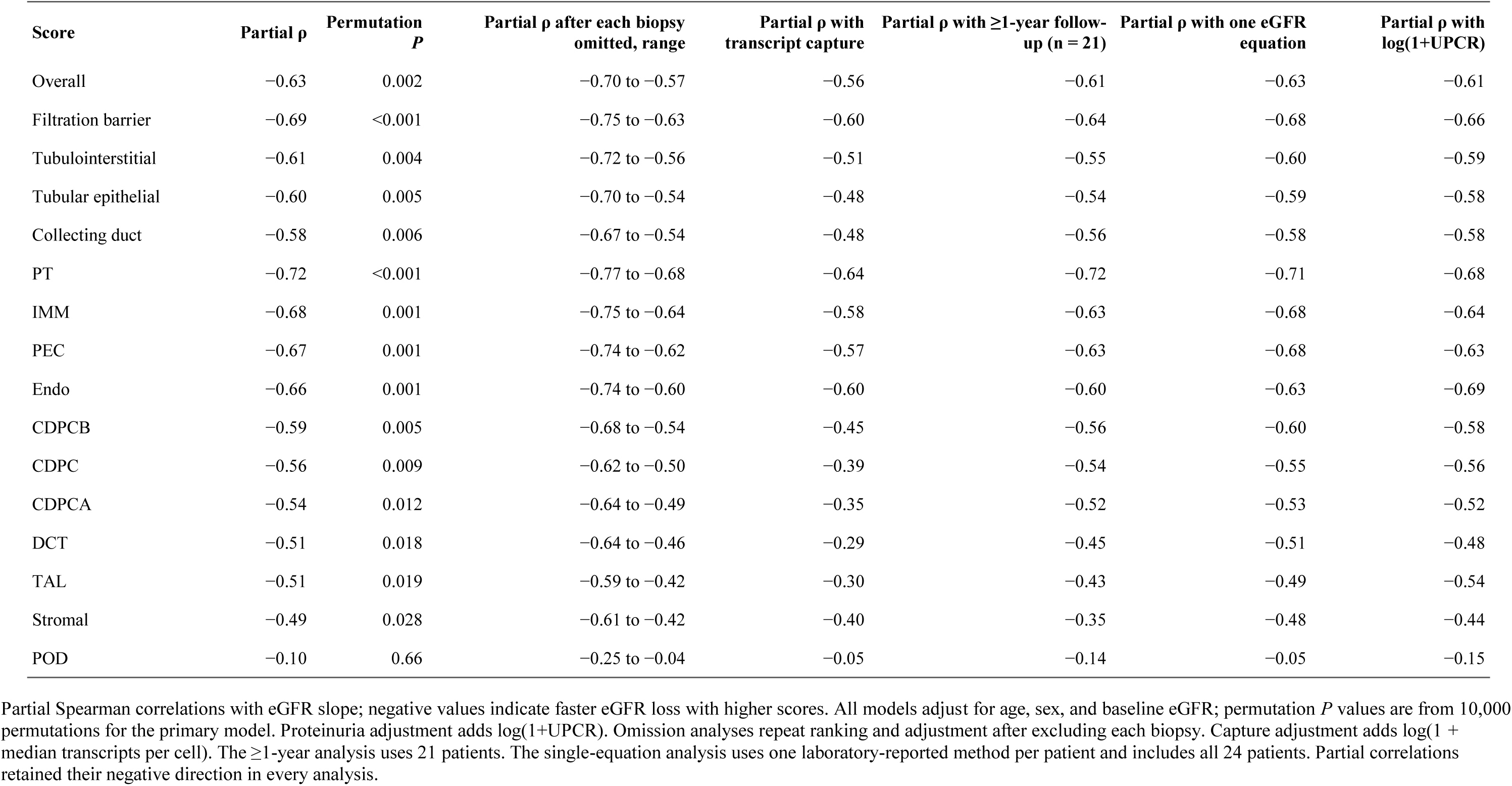
Tissue-aging scores and eGFR slope in 24 DKD patients: primary and sensitivity analyses.

| **Score** | **Partial ρ** | **Permutation *P*** | **Partial ρ after each biopsy omitted, range** | **Partial ρ with transcript capture** | **Partial ρ with ≥1-year follow-up (n = 21)** | **Partial ρ with one eGFR equation** | **Partial ρ with log(1+UPCR)** |
|---|---|---|---|---|---|---|---|
| Overall | −0.63 | 0.002 | −0.70 to −0.57 | −0.56 | −0.61 | −0.63 | −0.61 |
| Filtration barrier | −0.69 | <0.001 | −0.75 to −0.63 | −0.60 | −0.64 | −0.68 | −0.66 |
| Tubulointerstitial | −0.61 | 0.004 | −0.72 to −0.56 | −0.51 | −0.55 | −0.60 | −0.59 |
| Tubular epithelial | −0.60 | 0.005 | −0.70 to −0.54 | −0.48 | −0.54 | −0.59 | −0.58 |
| Collecting duct | −0.58 | 0.006 | −0.67 to −0.54 | −0.48 | −0.56 | −0.58 | −0.58 |
| PT | −0.72 | <0.001 | −0.77 to −0.68 | −0.64 | −0.72 | −0.71 | −0.68 |
| IMM | −0.68 | 0.001 | −0.75 to −0.64 | −0.58 | −0.63 | −0.68 | −0.64 |
| PEC | −0.67 | 0.001 | −0.74 to −0.62 | −0.57 | −0.63 | −0.68 | −0.63 |
| Endo | −0.66 | 0.001 | −0.74 to −0.60 | −0.60 | −0.60 | −0.63 | −0.69 |
| CDPCB | −0.59 | 0.005 | −0.68 to −0.54 | −0.45 | −0.56 | −0.60 | −0.58 |
| CDPC | −0.56 | 0.009 | −0.62 to −0.50 | −0.39 | −0.54 | −0.55 | −0.56 |
| CDPCA | −0.54 | 0.012 | −0.64 to −0.49 | −0.35 | −0.52 | −0.53 | −0.52 |
| DCT | −0.51 | 0.018 | −0.64 to −0.46 | −0.29 | −0.45 | −0.51 | −0.48 |
| TAL | −0.51 | 0.019 | −0.59 to −0.42 | −0.30 | −0.43 | −0.49 | −0.54 |
| Stromal | −0.49 | 0.028 | −0.61 to −0.42 | −0.40 | −0.35 | −0.48 | −0.44 |
| POD | −0.10 | 0.66 | −0.25 to −0.04 | −0.05 | −0.14 | −0.05 | −0.15 |

Partial Spearman correlations with eGFR slope; negative values indicate faster eGFR loss with higher scores. All models adjust for age, sex, and baseline eGFR; permutation *P* values are from 10,000 permutations for the primary model. Proteinuria adjustment adds log(1+UPCR). Omission analyses repeat ranking and adjustment after excluding each biopsy. Capture adjustment adds log(1 + median transcripts per cell). The ≥1-year analysis uses 21 patients. The single-equation analysis uses one laboratory-reported method per patient and includes all 24 patients. Partial correlations retained their negative direction in every analysis.

**Supplementary Table S4.**
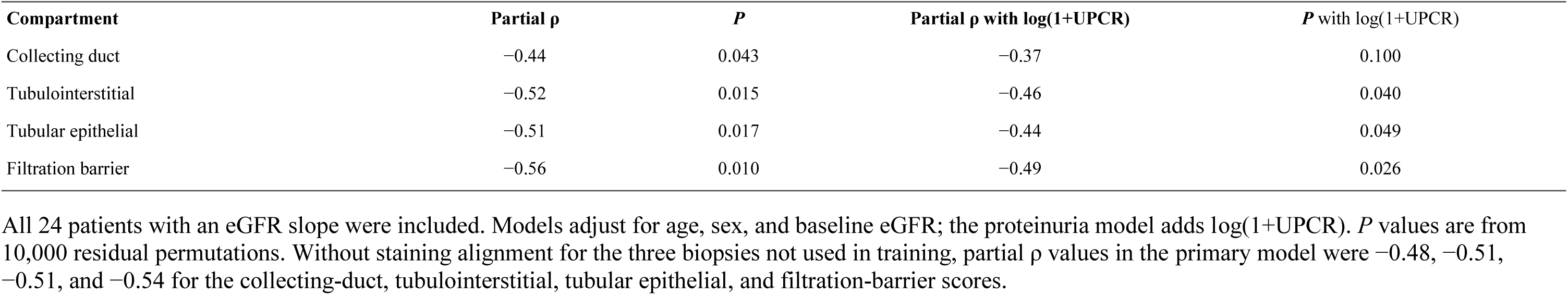
PAS-estimated compartment scores and eGFR slope with additional adjustment for proteinuria.

| **Compartment** | **Partial ρ** | ***P*** | **Partial ρ with log(1+UPCR)** | ***P*** with log(1+UPCR) |
|---|---|---|---|---|
| Collecting duct | −0.44 | 0.043 | −0.37 | 0.100 |
| Tubulointerstitial | −0.52 | 0.015 | −0.46 | 0.040 |
| Tubular epithelial | −0.51 | 0.017 | −0.44 | 0.049 |
| Filtration barrier | −0.56 | 0.010 | −0.49 | 0.026 |

All 24 patients with an eGFR slope were included. Models adjust for age, sex, and baseline eGFR; the proteinuria model adds log(1+UPCR). *P* values are from 10,000 residual permutations. Without staining alignment for the three biopsies not used in training, partial ρ values in the primary model were −0.48, −0.51, −0.51, and −0.54 for the collecting-duct, tubulointerstitial, tubular epithelial, and filtration-barrier scores.

## Supplementary methods

### Participants and tissue measurements

The analysis unit was one person; one control contributed two sections, which were combined. All 28 DKD biopsies had 11 cell-group scores, five overall/compartment scores, 66 pairwise proximity measurements, cell areas, and median transcript capture. Control proximity measurements were available for 23 participants.

### Control spatial measurements and spatial aging index

Neighbors of each cell were its six nearest cells by centroid distance. For control i and cell-pair e, observed neighbor count *O_ie_* was compared with expected count *E_ie_*:

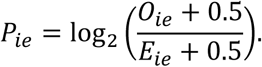

A negative age coefficient indicates lower observed-to-expected proximity in older tissue. The primary model was

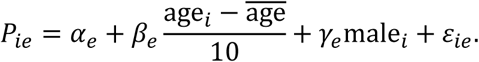

Four predefined summaries represented immune–epithelial, endothelial–podocyte, stromal–epithelial, and epithelial–epithelial relationships. Individual-pair models additionally included the fractions of the two participating cell types:

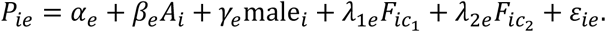

Same-group pairs used one fraction term. Benjamini–Hochberg correction was applied within measurement families; the four proximity-summary tests formed one family.

The spatial aging index combined selected, standardized control tissue measurements. For each control h, feature selection, scaling, and weighting were repeated using the other 23 controls, yielding an independently scored value 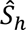. The age correlation was compared with 500 reassignments of control ages, each repeating this procedure:

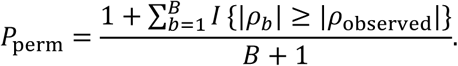

### Molecular tissue-aging scores

For biopsy i, cell group c, and gene g, the cell-group mean count was transformed as 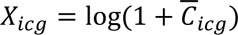. Candidate genes were detected in at least 15% of control profiles and passed the neighboring-cell-group specificity assessment. Within each group, the 20 smallest sex-adjusted age-model *P* values in each direction defined fixed gene sets 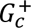 and 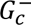:

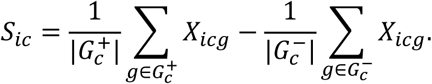

Gene selection was by rank rather than a whole-screen false-discovery threshold. Each cell-group score used all 20 genes in each direction. In control cells held out from model fitting, cell-group assignment had an overall accuracy of 0.884.

The collecting-duct score averaged CDPC, CDPCA, and CDPCB; tubular epithelial score averaged DCT, PT, and TAL; tubulointerstitial score averaged PT, TAL, DCT, and stromal cells; and filtration-barrier score averaged endothelium, podocytes, and parietal epithelial cells. The overall score averaged all 11 groups. All constituent groups were present in all 28 DKD biopsies. Spatial proximity, cell area, and transcript-capture measurements were not components of these molecular scores. Supplementary Table S1 lists all genes and control-age coefficients.

### PAS image processing and score estimation

Tissue was identified on a 32-fold downsampled thumbnail as pixels with grayscale intensity below 220 and saturation above 15. Tiles of 128 × 128 µm (486.92 pixels at 0.262877 µm per pixel) were placed on a non-overlapping grid, and tiles with at least 50% tissue were kept. Stain normalization followed Reinhard and colleagues [21] with an in-house implementation: tiles were converted from RGB to L*a*b* with OpenCV, and the mean and standard deviation of each channel were matched to a single target, the median channel statistics of up to 3,000 tiles sampled at random from all training slides, including those used for evaluation; the statistics were computed from tissue pixels only, excluding pixels with all three RGB channels above 200, and the transformation was applied to all pixels. Tiles were resized to 224 × 224 pixels and encoded with UNI2-h. Per-biopsy features were the 25th, 50th, and 75th percentiles and the mean across tiles (6,144 values). Within each training fold, scores were centered within each disease, features were reduced to 16 principal components, and a multi-output ridge regression with the penalty chosen by internal cross-validation estimated the four compartment scores. Folds were grouped by patient, the procedure was repeated 20 times with different fold assignments, and each training biopsy’s compartment estimate was the mean over the 20 repetitions. The four estimates were examined separately in the clinical analyses. Tile-level estimates were mapped to the original-slide pixel centres using the slide calibration of 0.262877 µm per pixel, with each tile extending 64 µm on either side of its centre. Maps illustrate variation within a biopsy; clinical inference used the patient-independent biopsy-level estimates (Supplementary Figure S6).

The three DKD biopsies not used in training were scored by the model fitted to all 48 training biopsies. Their slides were stained separately with stronger PAS staining, and their estimates were aligned by the mean difference between the two staining sets in IgA nephropathy biopsies, which agreed with the difference measured on paired sections from the same blocks; results were similar without this alignment (Supplementary Table S4).

To localize morphological contributions to the tile-level tissue-aging estimate, 16-µm image blocks were flattened in turn, and each modified tile was re-embedded and scored. A fall in the estimate after occlusion identified a region contributing to a higher estimate (red outline); a rise identified a region contributing to a lower estimate (blue outline). Representative control and DKD tiles were displayed from the lowest to the highest estimate quintile, and the highest-estimate examples from control, DKD, and IgA nephropathy were shown without and with outlines (Figure 6).

### Clinical measurements and eGFR trajectories

Clinical information was linked to each biopsy using specimen records, with clinical review for two records requiring manual linkage. Baseline eGFR was laboratory-reported, with the calculation method recorded as MDRD in 18 biopsies and EPI in 10. The exact equation versions were unavailable.

All laboratory eGFR records on or after biopsy were collapsed to one median per observed day before slope estimation. Where both reported methods occurred on one day, the daily median included both. For a patient with observation times *t_j_*, in years from biopsy, and daily eGFR *y_j_*, the eGFR slope was

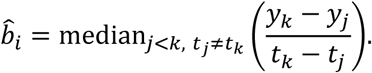

A primary trajectory required at least two distinct dates spanning at least 0.5 years. Four patients had observed spans shorter than 6 months; they were retained in the cohort and all baseline, molecular, and spatial analyses. All primary slope models used the remaining 24 patients.

Measurements after the onset of kidney failure were not used. Kidney failure was identified as eGFR values at or below 8 mL/min/1.73 m²sustained for at least 6 months, beginning at the first such value; in one patient who received a kidney transplant, measurements from transplantation onward were not used. Measurements during one hospitalization with acute kidney injury and temporary dialysis were also not used.

Baseline UPCR was available for 27 biopsies. For a missing baseline value, the first measured UPCR within 14 days after biopsy was accepted as a peri-biopsy measure. This was needed for one biopsy (measured 10 days after biopsy).

Additional analyses required at least one year between observed eGFR measurements or used one reported eGFR method per patient. For the latter, the method with the most distinct post-biopsy dates was selected before outcome testing, with EPI breaking a tie; slopes used daily medians from that method and the same six-month requirement. All 24 patients remained eligible.

### Clinical models, assay capture, and inference

The association of each tissue score with eGFR slope was summarized by partial Spearman correlation: the score and slope were ranked, each ranked variable was regressed on age, sex, and baseline eGFR, and the residuals were correlated. Two-sided *P* values were obtained from 10,000 permutations of the slope residuals.

Baseline eGFR and log(1+UPCR) models adjusted for age and sex; age-tracking models adjusted for sex. Clinical coefficients and intervals are reported for every predefined score.

Each biopsy was omitted once, repeating ranking, scaling, and model fitting.

The assay-capture sensitivity added standardized log(1+median transcripts per cell). All 24 slope-eligible biopsies had this measurement. A proteinuria sensitivity analysis added log(1+UPCR) to the same covariates.

Post-biopsy ACE inhibitor or angiotensin receptor blocker use was recorded for 26 patients, and SGLT2 inhibitor use for seven. These fields describe exposure sometime after biopsy, not baseline treatment or duration, and were not inserted as baseline covariates. Diabetes duration, glycated hemoglobin, and adequately timed treatment and blood-pressure measurements were not available for the fixed primary model.

PAS compartment estimates were related to eGFR slope in all 24 patients with both measurements, using the same age-, sex-, and baseline-eGFR-adjusted partial Spearman procedure as the transcriptomic scores. All four compartments were evaluated, with nominal two-sided permutation *P* values. Confidence intervals used 2,000 patient-bootstrap resamples, repeating ranking and covariate adjustment. A sensitivity analysis added log(1+UPCR).

### Spatial correspondence and disease context

Proximity summaries averaged eight immune–epithelial pairs, eight stromal–epithelial pairs, or the endothelial–podocyte pair.

Biopsy-level Spearman correlations related the overall molecular score to each proximity summary and median PEC or POD area, separately for controls and DKD. The DKD area analysis included all 28 biopsies. Models comparing DKD with controls across all 66 pairs adjusted for age and sex; control-age models adjusted for sex. The coefficient correlation was assessed against 4,999 control-age reassignments, preserving the covariance of the cell-pair measurements.

For expression, all 374 signature genes were excluded from every cell group, and genes detected in at least 15% of controls were tested. Control-age effects were estimated in controls with age and sex, DKD-versus-control effects in all participants with DKD status, age, and sex, and overall-score effects within DKD with the standardized score, age, and sex; all models included the log median transcripts per cell of the cell group. Effects were scaled to the control standard deviation. Correspondence was summarized as the Spearman correlation of gene effects within each cell group and across all groups, and permutation *P* values came from 1,000 permutations of control age or of the DKD score. Genes with a DKD-versus-control false discovery rate below 0.05 were divided into those that changed with control age in the same direction (*P* < 0.05) and the remainder, and each set was tested for Hallmark and Reactome overrepresentation against the genes tested in that cell group.

### Molecular programs and injured proximal tubule

Cell-group pseudobulk analyses included continuous control age with sex adjustment and an upper-versus-lower age-tertile contrast. Candidate gene-age associations were also assessed after accounting for the same gene in its highest-expressing cell group. Differential-expression reporting required within-cell-group *q* < 0.05, a same-direction tertile effect, and no compartment or sex-chromosome flag. These reporting criteria were separate from the selection by rank used for tissue-aging scores. Pathway analyses used Hallmark, Gene Ontology Biological Process, Reactome, and KEGG sets with tested Xenium panel genes as the universe. SenMayo, CellAge, and Fridman scores were tested at the level of the individual control.

The single-nucleus reference comprised 125 samples and 889,646 nuclei. Coexpression extended Xenium-derived programs to the full transcriptome but did not assess spatial proximity. For PT-state transfer, 124 reference samples with sufficient PT nuclei contributed 21,328 cells. The injured state comprised injured PT and PT-specific stressed epithelium. Reference counts were matched to Xenium PT depth, and classification was assessed in five partitions with sample separation. Shared-gene probabilities in Xenium PT cells were converted to percentile ranks within control-derived transcript-depth bins; biopsy burden was the mean percentile. DKD baseline and longitudinal PT-state models included sensitivity adjustment for log(1+median detected genes in PT).

Analyses used Python 3.14.6 (NumPy 2.4.4, pandas 3.0.2, SciPy 1.17.1, statsmodels 0.14.6) and R 4.5.3 (ggplot2 4.0.3, patchwork 1.3.2). Image analyses used Python 3.10.13 (PyTorch 2.13.0, timm 1.0.27, OpenSlide 4.0.1, OpenCV 4.10.0, and scikit-learn 1.7.2).

## Supplementary results

### Spatial and cellular changes in control kidneys

The continuous-age and upper-versus-lower-tertile analyses agreed in direction for the leading control relationships (Supplementary Figure S1). The spatial aging index evaluated with each control excluded gave ρ = 0.613, a bootstrap 95% interval of 0.19–0.84, and age-permutation *P* = 0.022. Cell fractions did not pass family-level correction.

### Molecular programs and senescence

Proximal-tubule and collecting-duct profiles involved oxidative phosphorylation and fatty-acid metabolism, whereas immune, endothelial, and stromal profiles involved interferon and adaptive-immune programs. SenMayo increased with age in podocytes (β = 0.011 per decade, *q* = 0.0026), rather than as a kidney-wide program. Cell-group pathway results are shown in Supplementary Figure S3.

The overall score correlated with median transcripts per cell at ρ = 0.10 in controls and 0.82 in DKD.

### Injured proximal tubule

In all 28 DKD biopsies, a 1-SD higher injured-PT burden corresponded to 16.6 mL/min/1.73 m²lower eGFR at biopsy (95% CI, 8.9–24.3 lower). The injured-PT burden showed no clear association with eGFR slope (Supplementary Figure S4).

### PAS-estimated scores

All four PAS-estimated compartment scores were inversely associated with eGFR slope in the 24 patients (Figure 5c), also with additional adjustment for log(1+UPCR) (partial ρ, −0.37 to −0.49) and without staining alignment for the three biopsies not used in training (−0.48 to −0.54; Supplementary Table S4).

Tile-level estimates mapped onto the DKD section in Figure 5a varied within the biopsy, and the four compartment maps showed similar spatial patterns (Supplementary Figure S6).

## Supplementary figure legends

**Supplementary Figure S1.**
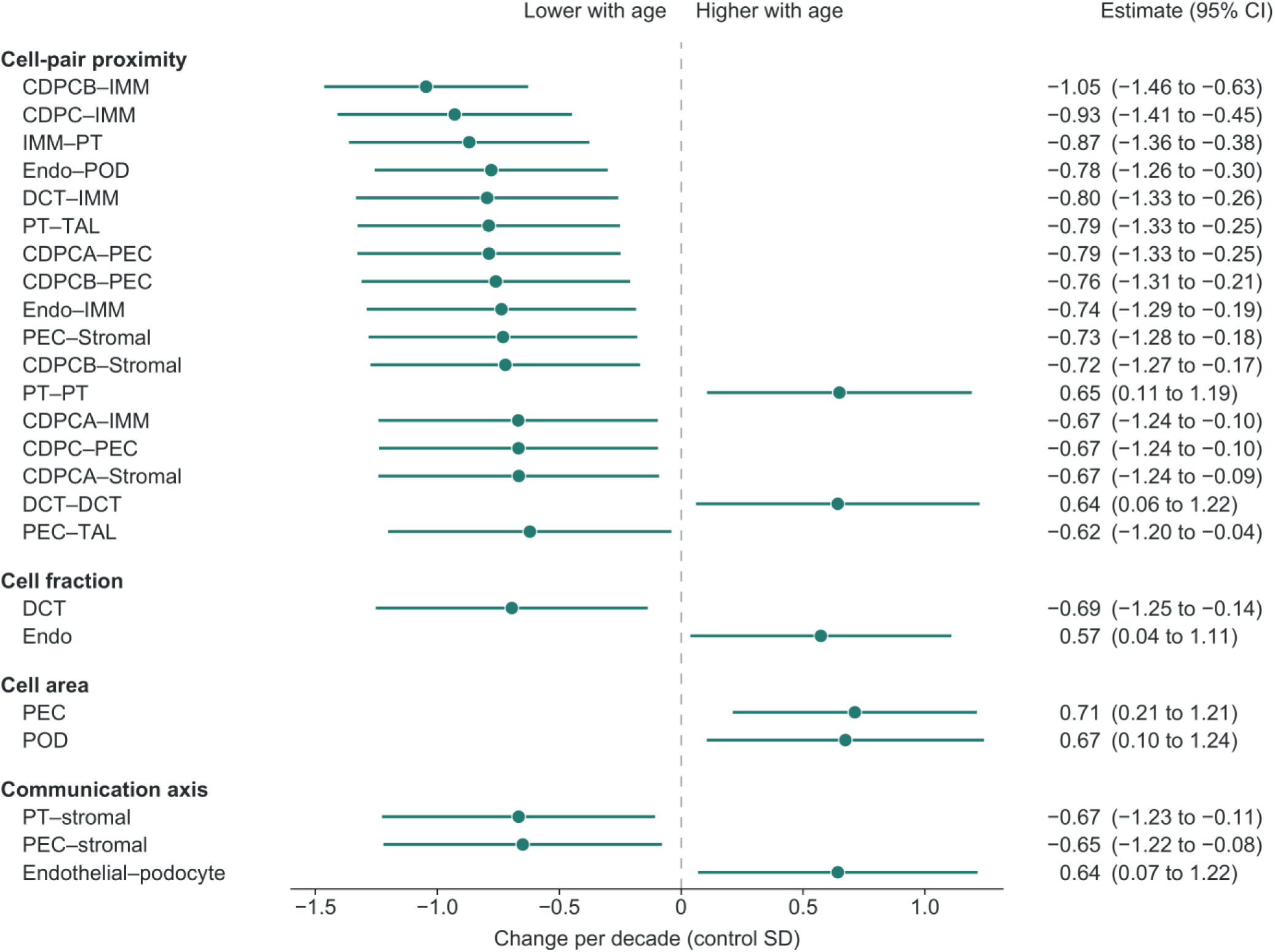
Control tissue measurements and chronological age. Leading sex-adjusted control-age associations across proximity, cell fraction, cell area, transcript content, ligand-receptor, and communication measurements. Effects and 95% confidence intervals are per decade of age, expressed in each measurement’s standard deviation among controls. The control, not the cell, is the analysis unit.

**Supplementary Figure S2.**
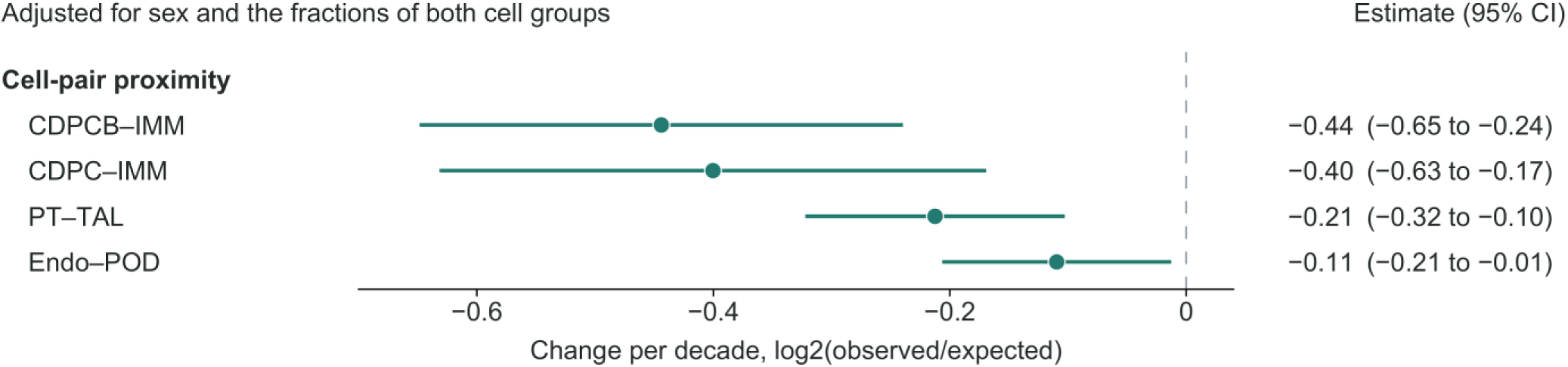
Cell-pair proximity and control age after adjustment for cell-group abundance. Change per decade in CDPCB–IMM, CDPC–IMM, PT–TAL, and Endo–POD proximity after adjustment for sex and the fractions of both cell groups, with 95% confidence intervals.

**Supplementary Figure S3.**
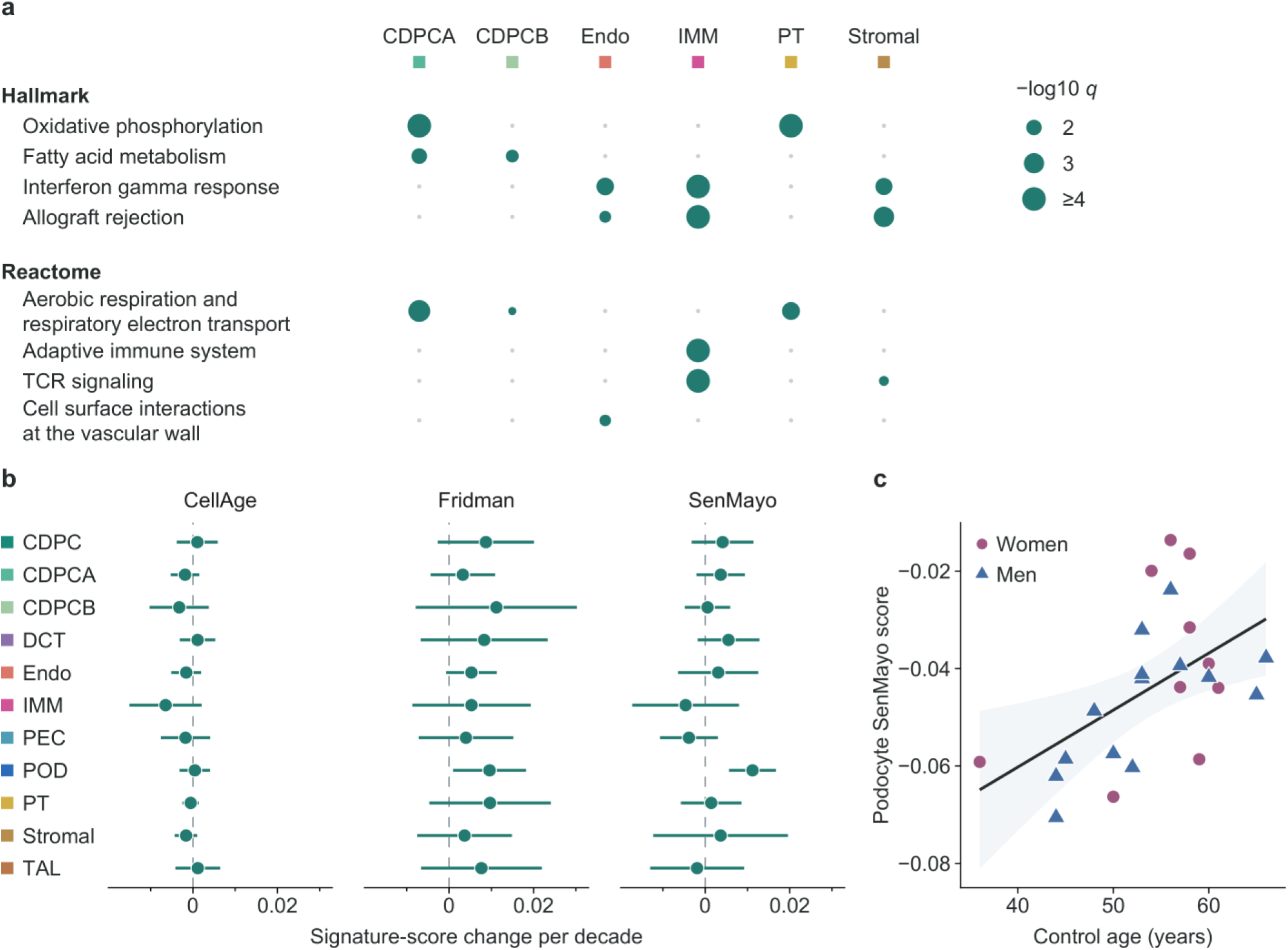
Cell-group expression and senescence programs across control age. a: Representative Hallmark and Reactome programs supported by the continuous-age and age-tertile analyses. b: Sex-adjusted per-decade changes in CellAge, Fridman, and SenMayo scores across cell groups, with 95% confidence intervals. c: Podocyte SenMayo score against control age; point color denotes sex.

**Supplementary Figure S4.**
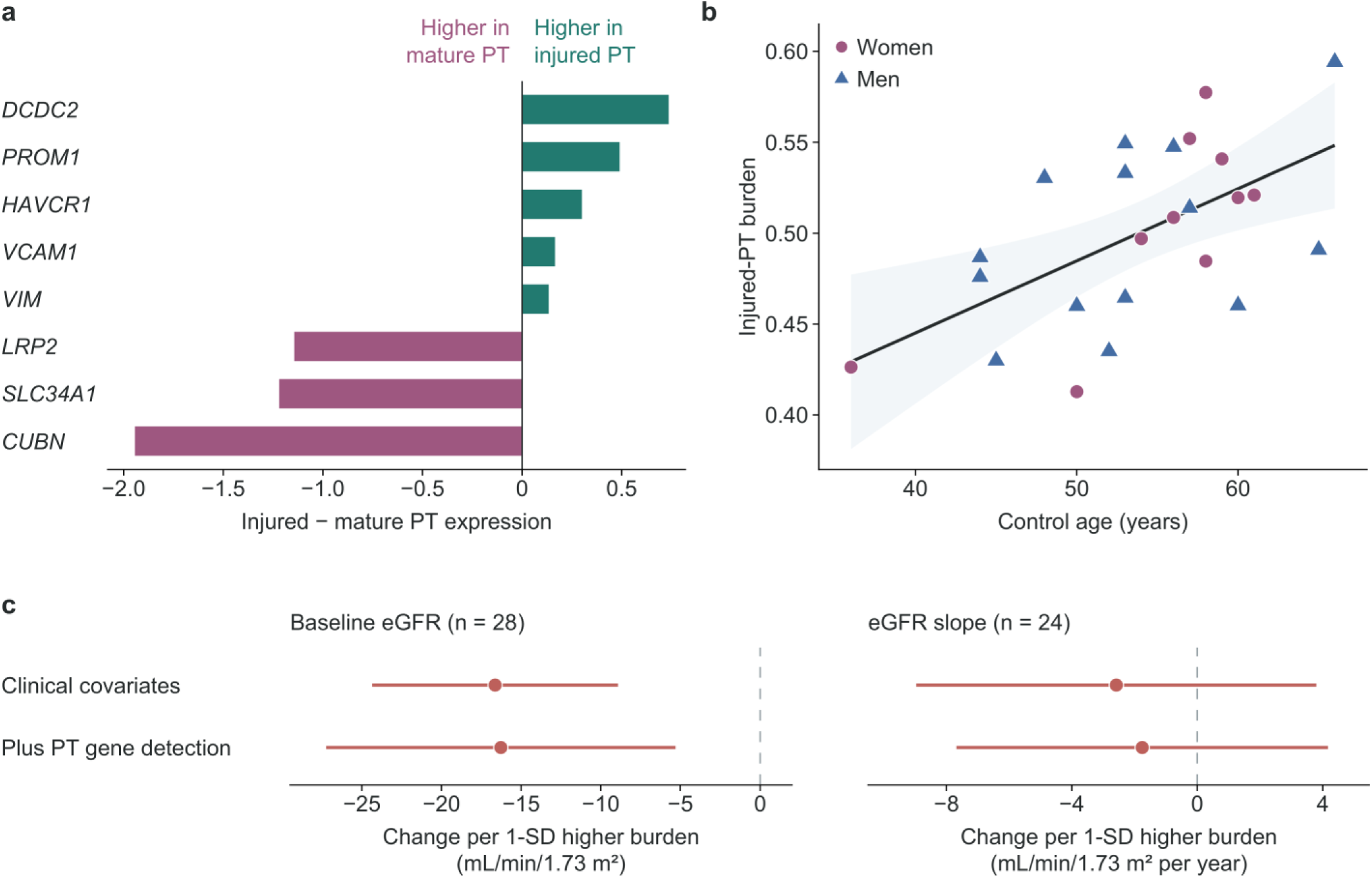
Injured proximal-tubule state defined in the single-nucleus reference. a: Expression differences between injured and mature PT for transport and stress markers in the reference. b: Control injured-PT burden against age, after calibration for transcript capture. c: Associations of injured-PT burden with baseline eGFR and eGFR slope in DKD, with and without further adjustment for detected genes in PT cells. Coefficients are per 1-SD higher burden; lines show HC3 95% confidence intervals.

**Supplementary Figure S5.**
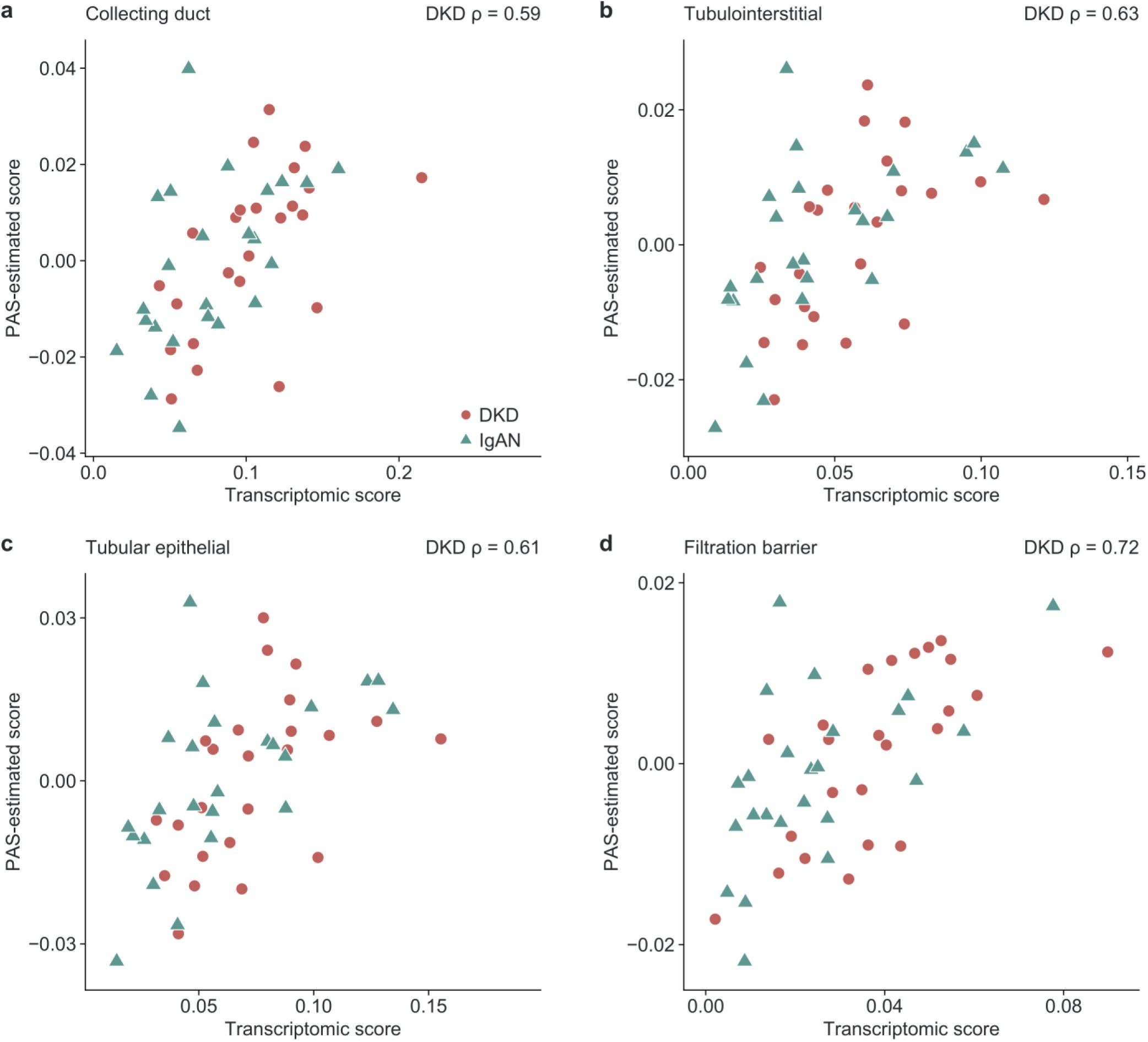
PAS-estimated against transcriptomic compartment scores. Scores estimated from PAS slides by models built without each patient, averaged over 20 cross-validation repetitions, are plotted against the transcriptomic collecting-duct, tubulointerstitial, tubular epithelial, and filtration-barrier scores in the 24 DKD and 24 IgA nephropathy biopsies used for training. Red circles denote DKD and jade triangles denote IgA nephropathy. Spearman ρ was computed in the DKD biopsies; axes are in the original score units.

**Supplementary Figure S6.**
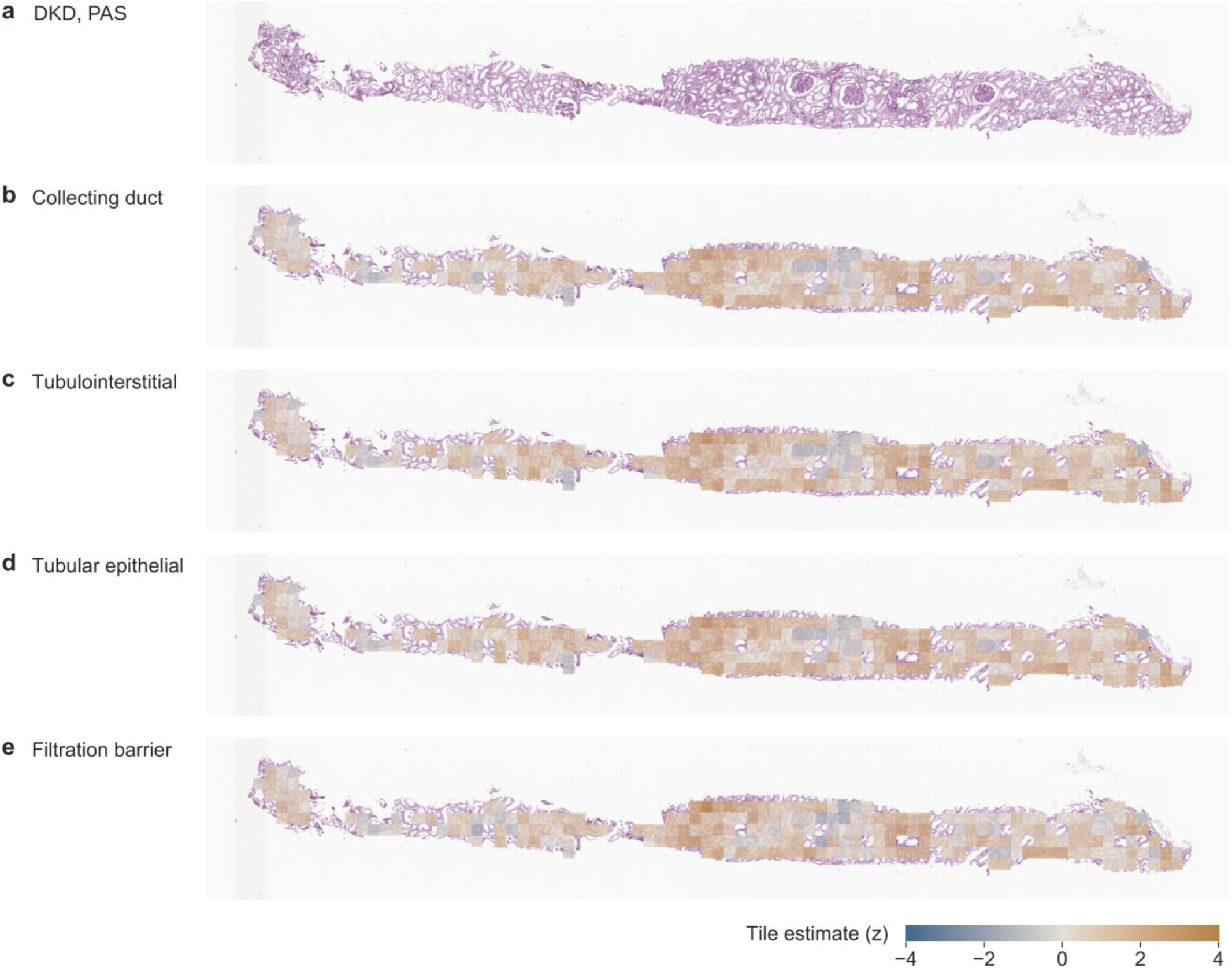
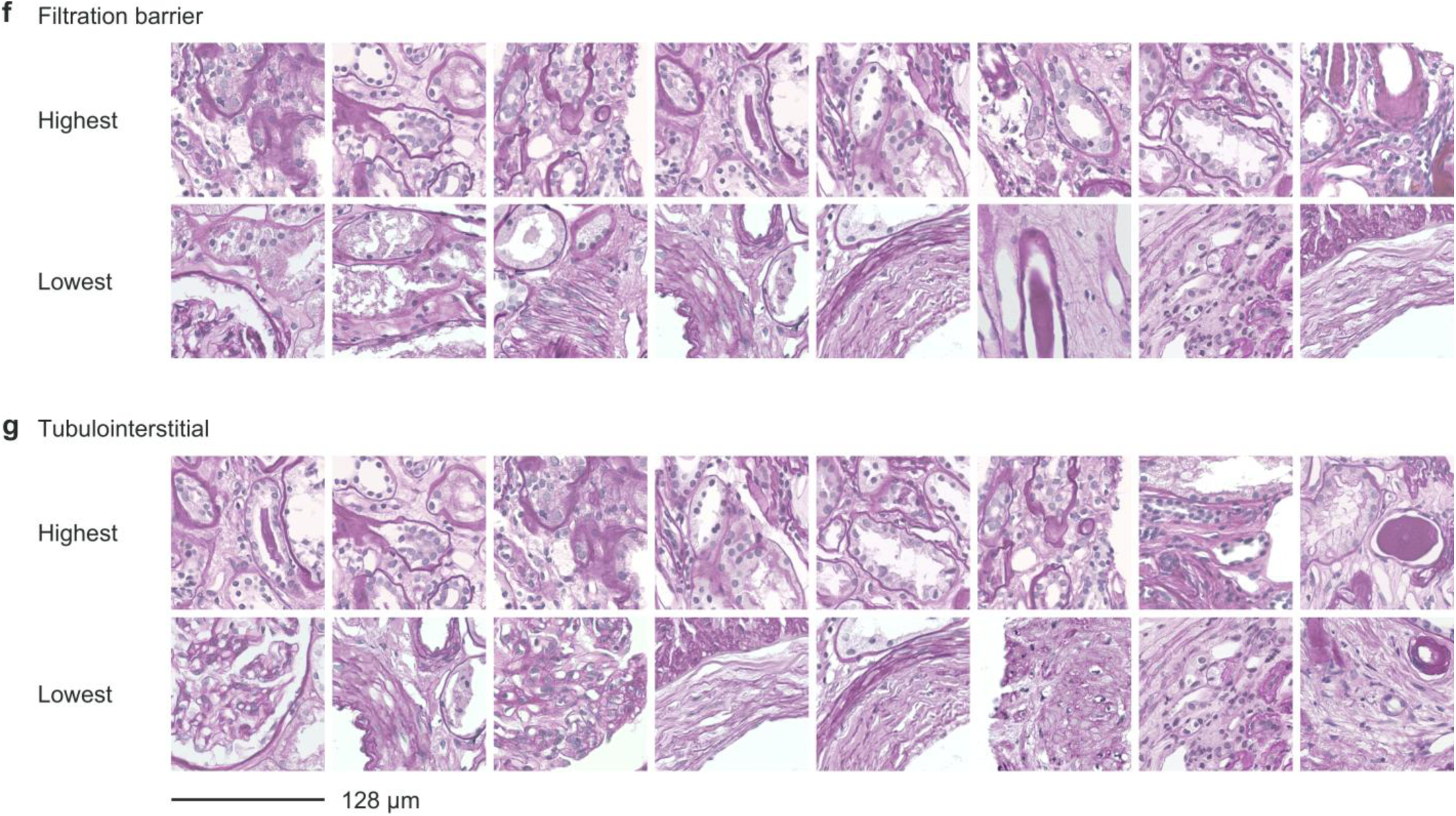
Tile-level PAS estimates. a–e: The DKD PAS section shown in Figure 5a (a) and the four compartment-specific tile maps (b–e). The section is rotated 90° for display, and each 128 × 128 µm tile is placed at its slide coordinates; the common color scale spans −4 to 4 standardized tile-score units. Tile-level estimates come from a model fitted to all 48 training biopsies and are shown for display only. f, g: Highest- and lowest-scoring 128 × 128 µm tiles for the filtration-barrier and tubulointerstitial scores, respectively, selected from the 24 slope-eligible DKD biopsies with at least 80% tissue coverage. Each row contains eight tiles; within each compartment, each biopsy contributes one tile across both rows. Tiles share a common stain-normalized display. Scale bar, 128 µm.

